# The ClinGen Severe Combined Immunodeficiency Disease Variant Curation Expert Panel: Specifications for classification of variants in *ADA*, *DCLRE1C*, *IL2RG*, *IL7R*, *JAK3*, *RAG1*, and *RAG2*

**DOI:** 10.1101/2025.02.11.25322033

**Authors:** Vanessa C. Jacovas, Michelle Zelnick, Shannon McNulty, Justyne E. Ross, Namrata Khurana, Xueyang Pan, Alejandro Nieto, Shiloh Martin, Benjamin McLean, Marwa A. Elnagheeb, Morton J. Cowan, Jennifer M. Puck, Mike Hershfield, James Verbsky, Jolan Walter, Eric Allenspach, Alice Y. Chan, Nicolai S. C. van Oers, Rajarshi Ghosh, Megan Piazza, Bo Yuan, Luigi D. Notarangelo, Britt A. Johnson, Ivan K. Chinn, the Severe Combined Immunodeficiency Variant Curation Expert Panel

## Abstract

**Purpose:** This collaborative study, led by the Clinical Genome Resource Severe Combined Immunodeficiency Disease Variant Curation Expert Panel (ClinGen SCID-VCEP), implemented and adapted the American College of Medical Genetics and Genomics/Association for Molecular Pathology (ACMG/AMP) guidelines for interpreting germline variants in genes with established relationships to SCID. The effort focused on the 7 most common SCID-related genes identified by SCID newborn screening in North America: *ADA*, *DCLRE1C*, *IL2RG*, *IL7R*, *JAK3*, *RAG1*, and *RAG2*.

**Methods:** The SCID-VCEP conducted a rigorous review of variants that involved database analyses, literature review, and expert feedback to derive gene-specific modifications to the ACMG/AMP guidelines. These specifications were validated using a pilot set of 90 variants. **Results:** Of these 90 variants, 25 were classified as pathogenic, 21 as likely pathogenic, 14 as variants of uncertain significance (VUS), 18 as likely benign, and 12 as benign. Seventeen variants with conflicting classifications in ClinVar were successfully resolved. The criteria included modifications to 20 of the 28 original ACMG/AMP criteria specific to SCID-related genes.

**Conclusion:** The SCID-specific variant curation guidelines developed by the SCID-VCEP will enhance the precision of SCID genetic diagnosis and provide a robust framework for interpreting variants in SCID-related genes, contributing to appropriate treatment of SCID.

## INTRODUCTION

Severe Combined Immunodeficiency Disease (SCID) represents a major clinical challenge due to severely compromised immunity that makes newborns extremely vulnerable to life-threatening infections^1^. Affecting approximately 1 in every 60,000 births in North America^2^, SCID requires early diagnosis and intervention for survival. Without timely and appropriate treatment, SCID is usually fatal within the first year of life. However, the advent of population-based newborn screening (NBS) now identifies SCID soon after birth, significantly improving the prospects for successful treatment ^3,4^. The 2022 International Union of Immunologic Societies (IUIS) classification of inborn errors of immunity summarized the genetic diversity of SCID, linking it to variants in 19 different genes and underscoring the complexity inherent to diagnosing and treating this condition^5^.

The treatment strategies for SCID are intricately linked to the genetic pathways affected, varying from enzyme replacement therapy to hematopoietic stem cell transplantation or gene therapy^6^. This genetic complexity necessitates genetic testing and precise interpretation for the development of optimal treatment plans. The introduction of advanced gene sequencing technologies has increased the challenge of classifying gene variants, thus making accurate curation a cornerstone in avoiding diagnostic errors and ensuring the deployment of suitable treatment pathways.

In response to this challenge, ClinGen^7^ has devised a framework to establish expert panels (EPs) tasked with refining the 2015 American College of Medical Genetics and Genomics/Association for Molecular Pathology (ACMG/AMP) sequence variant interpretation guidelines for specific gene-disease relationships^8^. Recognizing the pivotal role of accurate genetic diagnosis in SCID, the ClinGen Severe Combined Immunodeficiency Variant Curation Expert Panel (SCID-VCEP) was established in 2019. This expert panel is dedicated to adapting the ACMG/AMP guidelines for variant classification in the context of SCID, focusing first on seven genes that are most frequently implicated in SCID patients in North America and accounting for 80% of all reported cases captured by the Primary Immune Deficiency Treatment Consortium (PIDTC)^9^: *ADA* (adenosine deaminase, HGNC:186), *DCLRE1C* (DNA cross-link repair 1C, HGNC:17642), *IL2RG* (interleukin 2 receptor gamma subunit, HGNC:6010), *IL7R* (interleukin 7 receptor, HGNC:6024), *JAK3* (Janus kinase 3, HGNC:6193), *RAG1* (recombination activating gene 1, HGNC:9831), and *RAG2* (recombination activating gene 2, HGNC:9832)^10^.

This paper presents the culmination of the first efforts to refine variant classification processes to develop an improved curation framework for these critical genes. By submitting variant classifications to ClinVar as an “expert panel” submitter, resolving discrepancies, moving Variants of Uncertain Significance (VUS) towards Benign or Pathogenic status, and enhancing confidence in SCID variant classifications in ClinVar, this work marks a significant step forward in the genetic diagnosis and treatment of SCID, ultimately helping to improve patient care and management in this challenging field.

## MATERIAL AND METHODS

### Creation of the SCID-VCEP

The SCID-VCEP^11^ was established in November 2019 and is affiliated with the ClinGen Immunology Clinical Domain Working Group. The SCID-VCEP was assembled to include multidisciplinary experts who have expertise in inborn errors of immunity: immunologists, clinical and molecular geneticists, genetic counselors, research scientists, and variant curation experts. All EP members disclosed potential conflicts of interest as required. The VCEP scope of work was designated as genes with established relationships to a SCID phenotype, especially as listed in Table 1 of the IUIS document^5^, focusing initially on 7 genes: *ADA*, *DCLRE1C*, *IL7R*, *JAK3*, *RAG1*, *RAG2* (all autosomal recessive mode of inheritance, AR), and *IL2RG* (X-linked recessive mode of inheritance, XLR).

**Table 1.** Population allele frequency thresholds.

| Genes | Contribution to SCID (%) | BA1<br>Penetrance: 50%<br>Prevalence= 1/5,000<sup>16</sup><br>Allelic heterogeneity = 1 | BS1<br>Penetrance: 100%<br>Prevalence= 1/50,000<sup>14,15</sup><br>Allelic heterogeneity = 1 | PM2_Supporting | The most frequent pathogenic variant<sup>a</sup> |
| --- | --- | --- | --- | --- | --- |
| *ADA* | 12.8 | 0.00721 | 0.00161 | 0.000174 | c.956_960del |
| *DCLRE1C* | 3.2 | 0.00346 | 0.00078 | 0.000032 | c.571C>T |
| *IL2RG* | 30.7 | 0.01110 | 0.00249 | 0.000124 | NA |
| *IL7R* | 7.6 | 0.00566 | 0.00126 | 0.000041 | c.221+2T>G |
| *JAK3* | 5.2 | 0.00447 | 0.00100 | 0.000115 | c.1351C>T |
| *RAG1* | 19.2 | 0.00872 | 0.00195 | 0.000102 | NA |
| *RAG2* | 19.2 | 0.00872 | 0.00195 | 0.000058 | NA |

<sup>a</sup>The most frequently observed pathogenic variant with confirmed pathogenicity in the context of Severe Combined Immunodeficiency (SCID) according to the Variant Classification Expert Panel (VCEP) specifications. The transcripts used are: *ADA* (NM\_000022.4), *DCLRE1C* (NM\_001033855.3), *IL7R* (NM\_002185.5), and *JAK3* (NM\_000215.4).

### ACMG/AMP specifications

Adapting the ACMG/AMP sequence variant interpretation guidelines^8^ for the seven target genes in relation to SCID was systematically developed through monthly teleconferences. The SCID Variant Curation Expert Panel (VCEP) meticulously reviewed each criterion in the ACMG/AMP guidelines, leading to the establishment of seven sets of rules or specifications: one for each gene. The finalized specifications received endorsement from the Sequence Variant Interpretation (SVI) Working Group and are publicly accessible on the Criteria Specification Registry (CSpec)^12^. The ACMG/AMP specifications are updated periodically, so to find the most current information please visit https://cspec.genome.network.

### Validation and Pilot testing

The specifications for the seven genes were validated using 90 pilot variants distributed nearly equally across the seven genes. Variants were nominated by SCID-VCEP members from ClinVar and internal laboratory data, with the objective of including a balance of known/suspected pathogenic or benign variants (75.6%) and VUS or variants with conflicting/missing assertions (24.4%). Additionally, variants were selected to represent various variant types: missense (52.2%), nonsense and frameshift (21.1%), synonymous (13.4%), splicing (8.9%), and intronic (4.4%). All pilot variants were annotated using RefSeq IDs NM_000022.4 (*ADA*), NM_001033855.3 (*DCLRE1C*), NM_000206.3 (*IL2RG*), NM_002185.5 (*IL7R*), NM_000215.4 (*JAK3*), NM_000448.3 (*RAG1*), and NM_000536.4 (*RAG2*).

Two trained biocurators independently evaluated each variant according to the SCID-VCEP standards using the ClinGen Variant Curation Interface^13^ to record criteria and evidence.

Three American Board of Medical Genetics and Genomics (or foreign equivalent) certified geneticists and one immunologist expert in the gene of interest independently reviewed this evidence, with final decisions made by consensus. Following Step 4 approval from ClinGen, these variants were then deposited into ClinVar with a 3-star rating, indicating expert panel review.

## RESULTS AND DISCUSSION

This article presents the results of the SCID-VCEP refinement process for adapting the ACMG/AMP variant interpretation guidelines to analyze variants genes with established relationships to SCID. The resulting specifications for the classification of germline variants in *ADA*, *DCLRE1C*, *Il2RG*, *IL7R*, *JAK3*, *RAG1*, and *RAG2* were approved by the SVI Working Group (July 20, 2023) and serve as a framework to guide the assessment of variant pathogenicity in these genes.

Of the 28 original ACMG/AMP criteria, 8 (BP1, BP2, BP3, BP4, BP5, BP6, PP2, and PP5) were determined not applicable for these genes and were excluded from the framework.

Another 20 of the 28 original criteria required gene and/or disease-specific alterations (BA1, BP7, BS1, BS2, BS3, PM1, PM2, PM3, PM4, PM5, PM6, PP1, PP3, PP4, PS1, PS2, PS3, PS4, and PVS1). Only one criterion was approved without additional modifications (BS4). This section presents a comprehensive presentation of the criteria along with specific details.

### Modification of the ACMG/AMP criteria based on specific classes of criteria

#### Variant frequency and use of control populations (BA1, BS1, BS2, PM2, PS4)

SCID is a rare condition with variable prevalence across North America, ranging from 1 in 58,000 newborns (95% CI: 1 in 46,000–80,000)^14^ to 1 in 65,000 (95% CI: 1 in 51,000-90,000)^15^. However, its incidence markedly increases in certain ethnic groups, particularly in the Middle East due to the pronounced population structure in these highly endogamous populations, reaching 1 in 5,000 live births^14,16^.

To establish stringent requirements for BA1 (allele frequency is greater than expected for disorder, Stand-alone), we used the highest reported prevalence of 1 in 5,000 in the Middle Eastern population^16^. For BS1 (allele frequency is greater than expected for disorder), we utilized a general number for the North American population and defined it as 1 in 50,000^14,15^. The penetrance, prevalence, and allelic/genetic heterogeneity used for each gene are displayed in Table 1 and were calculated using the CardioDB metrics allele frequency web tool^17^.

The SCID-VCEP recommends employing the Genome Aggregation Database (gnomAD) Grpmax Filtering Allele Frequency to assess BA1, BS1, and PM2; if unavailable, it should be based on the highest Minor Allele Frequency (MAF) considering all non-bottleneck populations. Given the absence of distinctly higher SCID incidence in any gnomAD bottleneck populations, as for example Ashkenazi Jewish and Finnish, we enabled the assessment of Allele Frequency in these populations for BS1.

Within our framework, the PM2 criterion (absent from population databases), which provides evidence for variant pathogenicity based on absent or extremely rare minor allele frequency, was downgraded to supporting strength per SVI Working Group guidelines^18^. For *ADA*, *DCLRE1C*, *IL7R*, and *JAK3,* the thresholds were based on the most frequent pathogenic variant present in gnomAD, excluding known founder variants. The pathogenic classifications were derived using other SCID-VCEP specifications while excluding PM2 as a criterion to avoid circularity. Regarding *RAG1* and *RAG2*, which both comprise a single exon, achieving the pathogenic classification in absence of PM2 proved challenging without applying PVS1 at its maximum strength. Furthermore, for *IL2RG*, only one likely pathogenic variant was listed in gnomAD, and it did not reach the pathogenic level using SCID-VCEP specifications. So, for these three genes, *RAG1*, *RAG2*, and *IL2RG*, we applied a second approach based on CardioDB calculations by reducing the allelic heterogeneity parameter. Table 1 contains all thresholds and settings. Additionally, to apply PM2, the variant cannot be observed in the homozygous state within any population.

Criterion PS4 (prevalence of the variant in affected individuals is significantly increased compared with the prevalence in controls) is based on the significantly higher prevalence of a variant in case cohorts versus control cohorts, which is considered strong evidence for pathogenicity. Ideally, published case-control studies are used as evidence. Most pathogenic or likely pathogenic (P/LP) variants in these genes are rare, and since limited case-control data are available for most variants, the VCEP recommends counting individual cases toward PS4. This PS4 criterion is specifically applied to *IL2RG*, which is XLR. Since the other 6 genes are AR, the PM3 criterion is utilized to count probands (see PM3 section below). To apply the PS4 code for *IL2RG*, the proband must meet SCID diagnostic criteria according to the PIDTC definitions for SCID^1^ (Table S1) and the variant must be sufficiently rare (meet PM2 criteria). The *IL2RG* PS4 specification has a sliding weight scale to account for the number of unrelated probands who meet the SCID diagnostic criteria. PS4 is applied with Very Strong strength when described in ≥4 unrelated probands, default Strong strength with 3 probands, Moderate strength with 2 probands, and Supporting strength with 1 proband. To avoid double counting of probands, the proband evaluated for PP4 is excluded from PS4 counting.

The BS2 criterion (observed in a healthy adult individual with full penetrance expected at anearly age) assesses the presence of a particular variant in a healthy adult, where full penetrance of the associated condition is expected at an early age. Identifying a variant in the homozygous state within a healthy adult provides strong evidence supporting a benign interpretation. Given that SCID is often diagnosed at birth through NBS programs, we tailored the application of this criterion to align with the inheritance pattern of the gene. For AR genes (*ADA*, *DCLRE1C*, *IL7R*, *JAK3*, *RAG1*, and *RAG2*), the BS2 criterion is applied at a supporting level when at least one unaffected adult individual is documented in the literature or population databases, such as gnomAD, carrying the variant in the homozygous state. For the *IL2RG* gene, which is XLR, to apply BS2 at the supporting level, the variant must be observed in at least two unaffected hemizygous individuals. BS2 may be applied at the strong level if a variant is observed in three or more unaffected individuals.

#### Functional data/Experimental (BS3, PS3)

For PS3 (well-established *in vitro* or *in vivo* functional studies support a damaging effect on the gene or gene product) and BS3 (well-established *in vitro* or *in vivo* functional studies show no damaging effect on protein function or splicing), we adopted the system previously described^19^ to set specific minimum quality standards for *in vitro* functional assays.

The VCEP first conducted literature reviews and identified the most prevalent categories of functional assays used for these genes: enzyme activity, recombination and repair activity, phosphorylation and binding assays, surface expression, and interaction profiling (Supplemental References 1-20). For each assay, a review of the literature was employed to define quality criteria by which the strength of the evidence provided could be assessed, including the number of basic controls, technical replicates, positive controls, and negative controls (See Supplemental Material 1).

The BS3 criterion is exclusively applied to *ADA*, based on the high specificity of enzymatic measurement of ADA activity. Conversely, for *IL2RG*, *IL7R*, *JAK3*, *DCLRE1C*, *RAG1*, and *RAG2* only PS3 is applicable. Additionally, the PS3 criterion may be applied at a default strength level of ‘strong’ when supported by data from an animal model that expresses the variant in question and recapitulates the SCID phenotype. Table 2 provides a summary of the assays utilized and the requisite strength of evidence for each gene, as determined by these criteria.

**Table 2.**
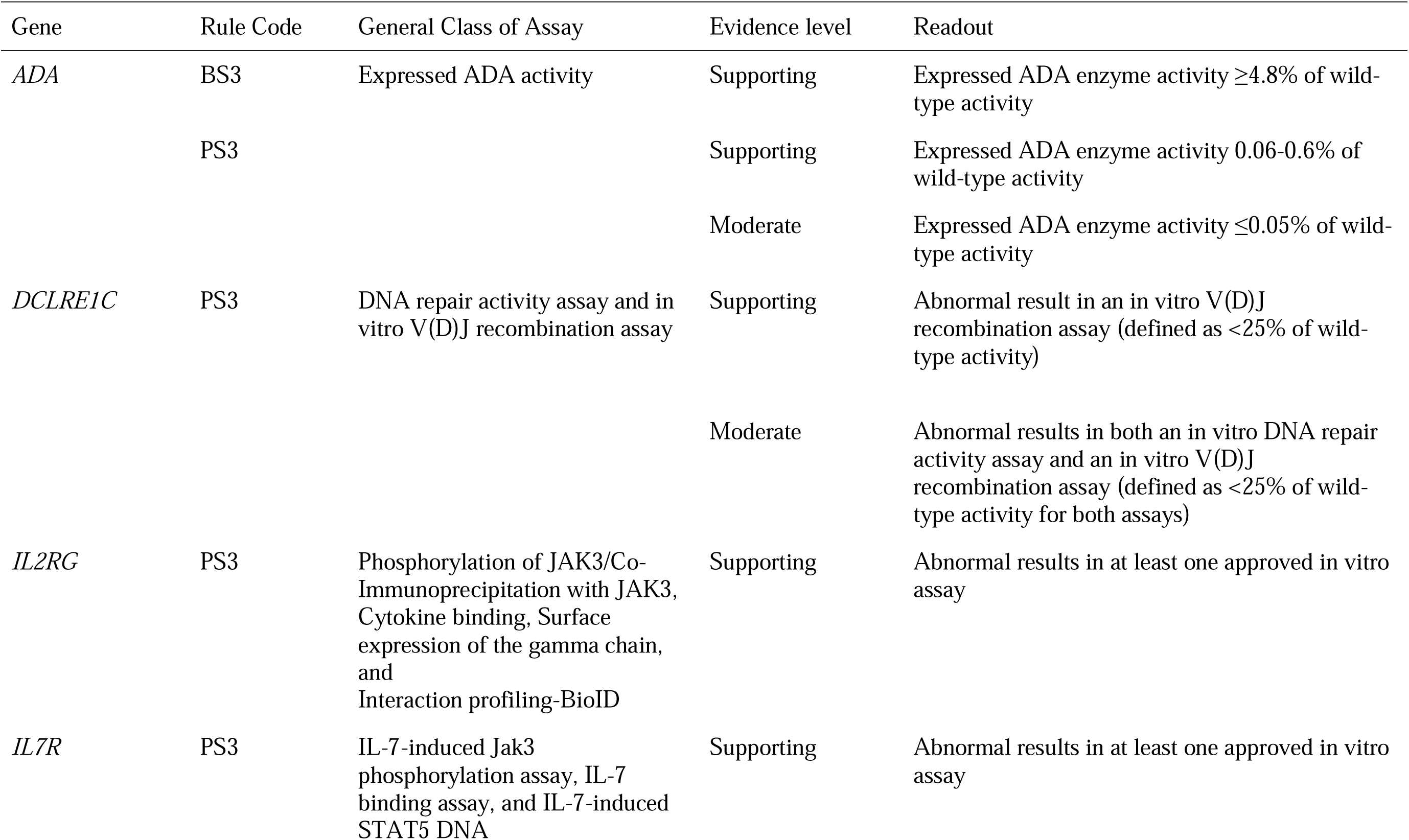
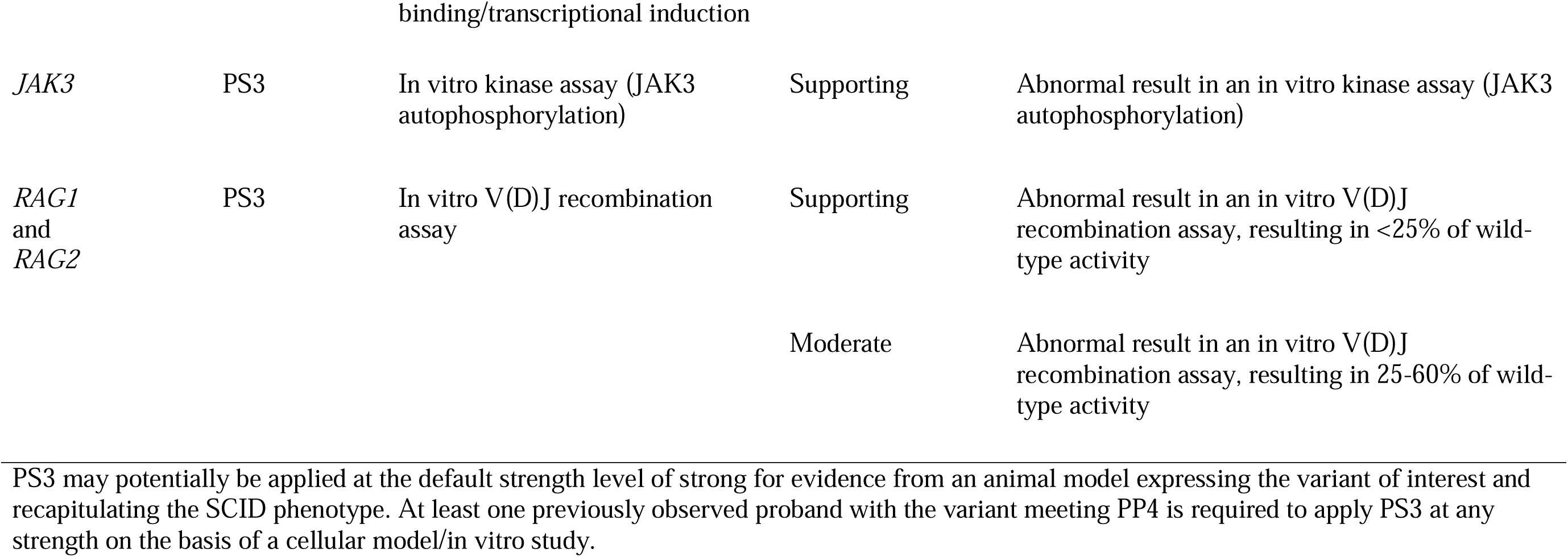
Functional assays (PS3/BS3) approved and strength of evidence:

| Gene | Rule Code | General Class of Assay | Evidence level | Readout |
| --- | --- | --- | --- | --- |
| *ADA* | BS3 | Expressed ADA activity | Supporting | Expressed ADA enzyme activity $\geq 4.8\%$ of wild-type activity |
|  | PS3 |  | Supporting | Expressed ADA enzyme activity 0.06-0.6% of wild-type activity |
| | | | Moderate | Expressed ADA enzyme activity $\leq 0.05\%$ of wild-type activity |
| *DCLRE1C* | PS3 | DNA repair activity assay and in vitro V(D)J recombination assay | Supporting | Abnormal result in an in vitro V(D)J recombination assay (defined as $<25\%$ of wild-type activity) |
| | | | Moderate | Abnormal results in both an in vitro DNA repair activity assay and an in vitro V(D)J recombination assay (defined as $<25\%$ of wild-type activity for both assays) |
| *IL2RG* | PS3 | Phosphorylation of JAK3/Co-Immunoprecipitation with JAK3, Cytokine binding, Surface expression of the gamma chain, and Interaction profiling-BioID | Supporting | Abnormal results in at least one approved in vitro assay |
| *IL7R* | PS3 | IL-7-induced Jak3 phosphorylation assay, IL-7 binding assay, and IL-7-induced STAT5 DNA | Supporting | Abnormal results in at least one approved in vitro assay |
| *JAK3* | PS3 | In vitro kinase assay (JAK3 autophosphorylation) | Supporting | Abnormal result in an in vitro kinase assay (JAK3 autophosphorylation) |
| *RAG1*<br>and<br>*RAG2* | PS3 | In vitro V(D)J recombination assay | Supporting | Abnormal result in an in vitro V(D)J recombination assay, resulting in <25% of wild-type activity |
|  |  |  | Moderate | Abnormal result in an in vitro V(D)J recombination assay, resulting in 25-60% of wild-type activity |

PS3 may potentially be applied at the default strength level of strong for evidence from an animal model expressing the variant of interest and recapitulating the SCID phenotype. At least one previously observed proband with the variant meeting PP4 is required to apply PS3 at any strength on the basis of a cellular model/in vitro study.

#### Critical domain (PM1)

PM1 (mutational hot spots and well-established functional domains) is defined by the SCID- VCEP for use at varying strength levels, from supporting to strong, depending on the gene and the region affected. Well-established functional domains were determined based on the known functions of the proteins and basic research defining these domains. For *ADA*, *DCLRE1C*, and *IL7R*, PM1 is not applicable due to the absence of such regions in these genes. For *IL2RG*, *JAK3*, *RAG1*, and *RAG2*, the relevant regions and the strength of their applicability are detailed in Table 3.

**Table 3.**
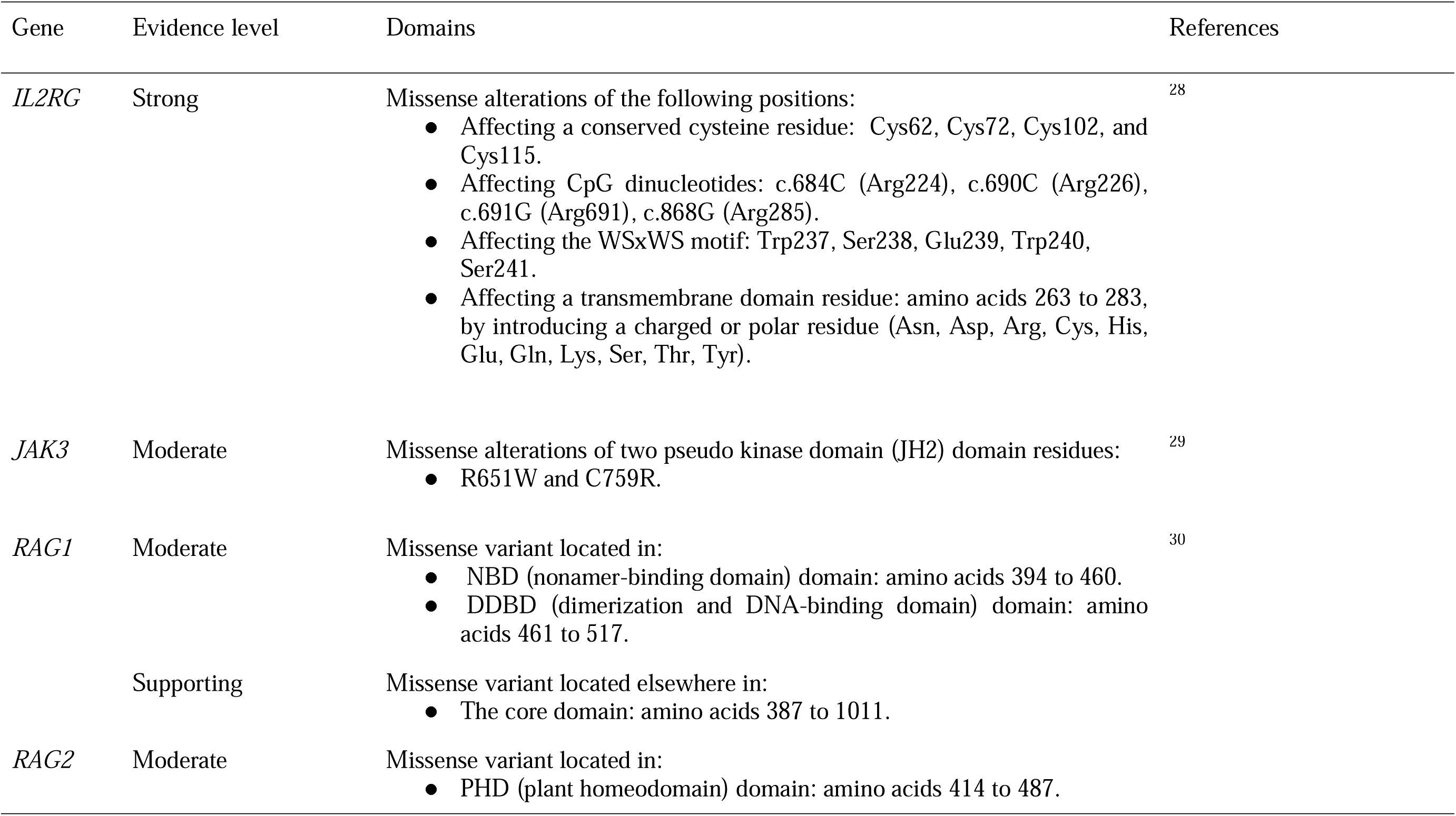
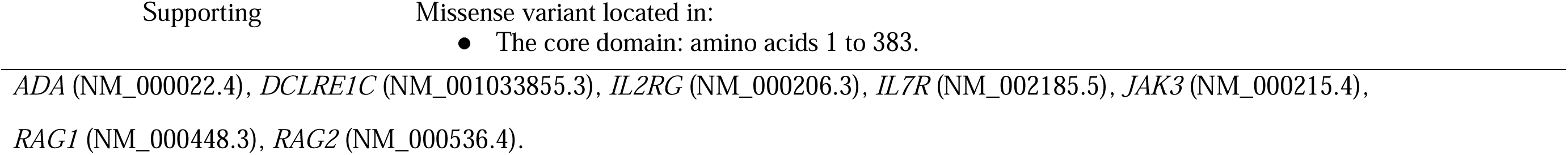
Critical Domains for Genes (PM1): Strength of Evidence.

#### Computational and predictive data (PVS1, PM4, PP3, BP7, PM5, PS1)

The PVS1 criterion (predicted null variant in a gene where loss of function is a known mechanism of disease) evaluates the impact of loss-of-function variants on gene function and potential pathogenicity. The level of evidence it provides towards pathogenicity can range from ‘very strong’ to ‘supporting,’ depending on where in the gene the variant occurs. The ClinGen SVI Working Group has provided detailed guidelines for how to apply this criterion effectively^20^. In accordance with the published PVS1 flowchart, we specified PVS1 on the basis of well-established evidence for our seven genes - based on their specific characteristics and genomic locations. The flowcharts depicting all types of loss-of-function variants and specific gene adaptations can be found in Supplemental Material 2.

Additionally, the PM4 criterion is applicable to in-frame deletions or insertions smaller than an entire exon, as well as in-frame whole-exon duplications that do not meet the PVS1 criteria. For PM4 to be applied at its default strength to deletion variants, the affected region must encompass a variant that has been previously established as pathogenic or likely pathogenic without predictions or observations of altered splicing. Conversely, if the region contains a VUS that is also not predicted or observed to affect splicing, then PM4 may be applied at a supporting level, denoted as PM4_Supporting.

The SCID-VCEP decided not to utilize *in silico* predictors for the PP3 criterion when evaluating missense variants due to insufficient validated evidence regarding the efficacy of these tools in this specific gene set. The PP3 criterion may still be applied for synonymous or intronic variants that are predicted by SpliceAI to affect splicing, provided they exhibit a delta score greater than or equal to 0.2.

The group agreed to adopt BP7, as delineated by ACMG/AMP, for synonymous variants where splicing prediction algorithms anticipate no effect and to broaden the criterion to encompass intronic variants situated at or beyond the +7 or -21 nucleotide positions. For a variant to meet BP7, it must be forecasted by a minimum of two out of three computational tools to have no influence on splicing. Considering the possible low conservation of nucleotides across vertebrate genes associated with T cell and B cell development, the presence of nucleotide conservation is not a prerequisite for the application of BP7.

In the context of variants impacting the same amino acid (AA) residue, the SCID-VCEP refined the application of the ACMG/AMP criteria by introducing nuanced recommendations for two rules: PS1 and PM5. For PS1 (same AA change as a previously established pathogenic variant), a new strength level of PS1_moderate was introduced for changes corresponding to a variant deemed likely pathogenic. Conversely, the PM5 criterion is applied with supporting strength (PM5_Supporting) when a different missense change at the same residue has been previously classified as likely pathogenic. The previously established variant must have been designated as pathogenic or likely pathogenic using SCID-VCEP specific standards.

### Case/Segregation

#### Segregation data (PP1 and BS4)

PP1 (co-segregation with disease in multiple affected family members) is applied according to the guidelines developed by the ClinGen Hearing Loss VCEP in collaboration with the SVI. This approach considers the logarithm of the odds (LOD) score and the number of affected and unaffected segregations to determine whether PP1 can be used at a supporting, moderate, or strong level^21^. A LOD score of ≥0.6 to <1.2 is required to use PP1 at a supporting level, a score of ≥1.2 to <1.5 for a moderate level, and a score of ≥1.5 for a strong level.

BS4 (Lack of segregation in affected members of a family) can be applied without additional specifications. It is used when a proband is phenotype-positive and genotype-negative.

#### De novo data (PM6 and PS2)

In SCID patients, *de novo* variants in AR genes are relatively rare, as with other AR disorders. However, occurrences have been reported^22^. Additionally, *de novo* variants in patients with *IL2RG*-associated SCID are more common^23^. For these cases, the SCID-VCEP specified the PM6 *de novo* (maternity and paternity not confirmed) and PS2 *de novo* (maternity and paternity confirmed) criteria following SVI recommendations (Table S2)^18^. The SVI-recommended approach bases the strength level for PM6/PS2 on confirmed versus assumed maternity/paternity status, the number of *de novo* probands, and phenotypic consistency. To achieve the highest level of phenotypic consistency (“phenotype highly specific for gene”), the patient must meet at least the PP4_Moderate criteria. For “phenotype consistent with the gene but not highly specific,” the proband must meet PP4. For “phenotype consistent with gene but not highly specific and high genetic heterogeneity,” the proband must exhibit a SCID phenotype without meeting PP4 criteria.

#### Variant phasing (PM3)

For the utilization of PM3 (detected *in trans* to a pathogenic variant), the SCID-VCEP followed SVI guidance for points per proband^18^. This guidance assigns greater point values to probands carrying a pathogenic or likely pathogenic variant confirmed *in trans* by either parental testing or cloning assays. In contrast, suspected in *trans* or homozygous occurrences are assigned lower point values (Table S3). Additionally, the application of PM3 requires that both variants (or the homozygous variant) in the proband occur at sufficiently rare allele frequencies that they meet the PM2 threshold. The applicability of PM3 to suspected founder variants with allele frequencies exceeding the PM2 threshold is evaluated on a case-by-case basis by the VCEP.

#### Phenotype (PP4)

Recently, PIDTC published updated guidelines for SCID diagnostic criteria, which we deployed for PP4 (patient’s phenotype or family history is highly specific for a disease with a single genetic etiology) assessments^1^. Due to specificities in the etiology of SCID, according to the gene/pathway affected, in Table 4 we present the details for PP4 according to each gene. The strength of PP4 applicability is determined by the total points accumulated by the most specific patient in the literature carrying the variant. If the score is less than 1 point, PP4 is not applied at any strength. For scores from 1 to less than 2 points, PP4 is met at the default level (Supporting). For moderate and strong strengths, the requirements vary by gene: For *ADA*: 2 to <9 points: PP4_Moderate; ≥9 points: PP4_Strong; *IL2RG*: ≥2 to <7 points: PP4_Moderate; ≥8 points: PP4_Strong; *DCLRE1C*: 2 to <7 points: PP4_Moderate; ≥7 points: PP4_Strong; *IL7R* and *JAK3*: 2 to <6 points: PP4_Moderate; ≥6 points: PP4_Strong; *RAG1* and *RAG2*: ≥2 to <4 points: PP4_Moderate; ≥4 points: PP4_Strong.

**Table 4.**
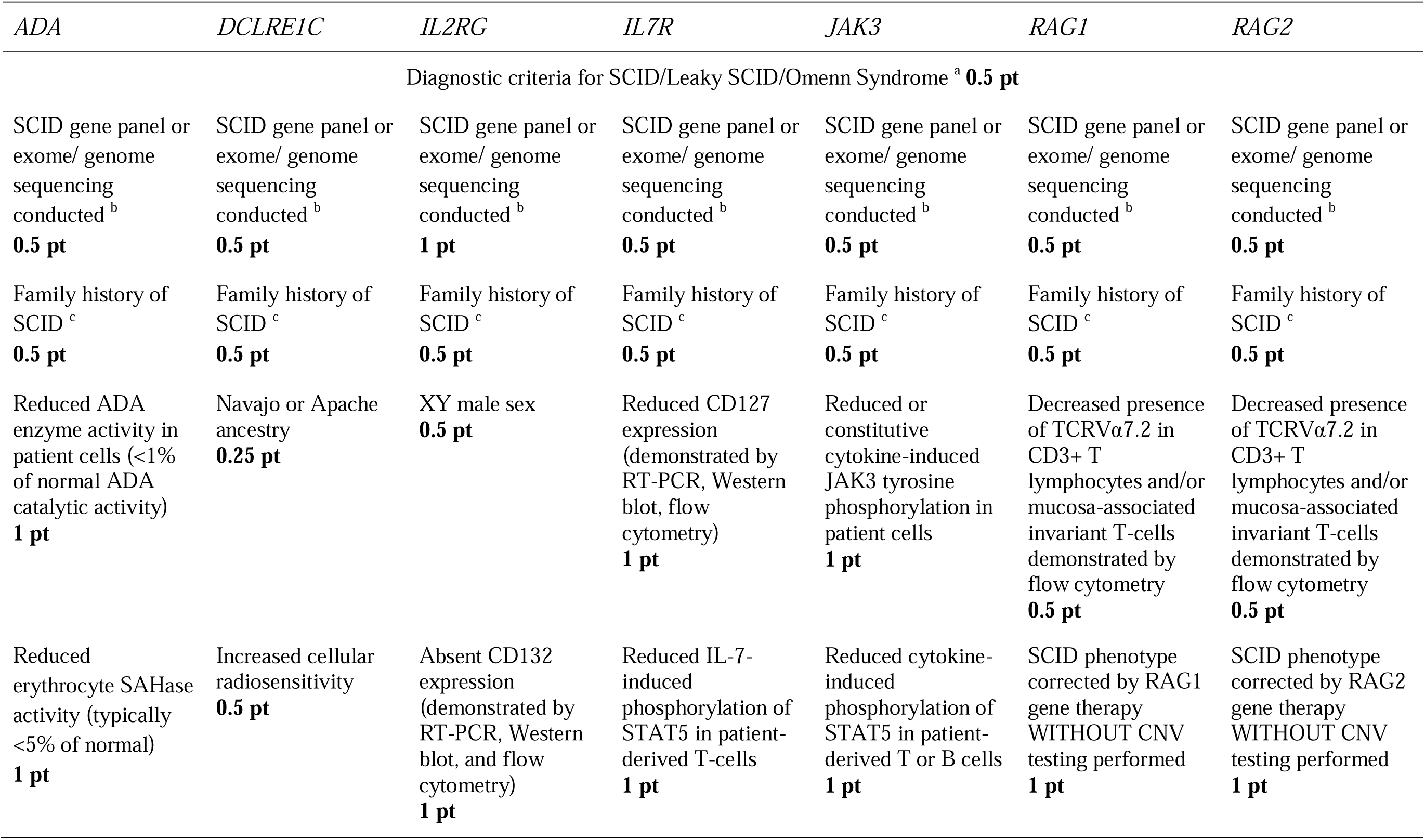
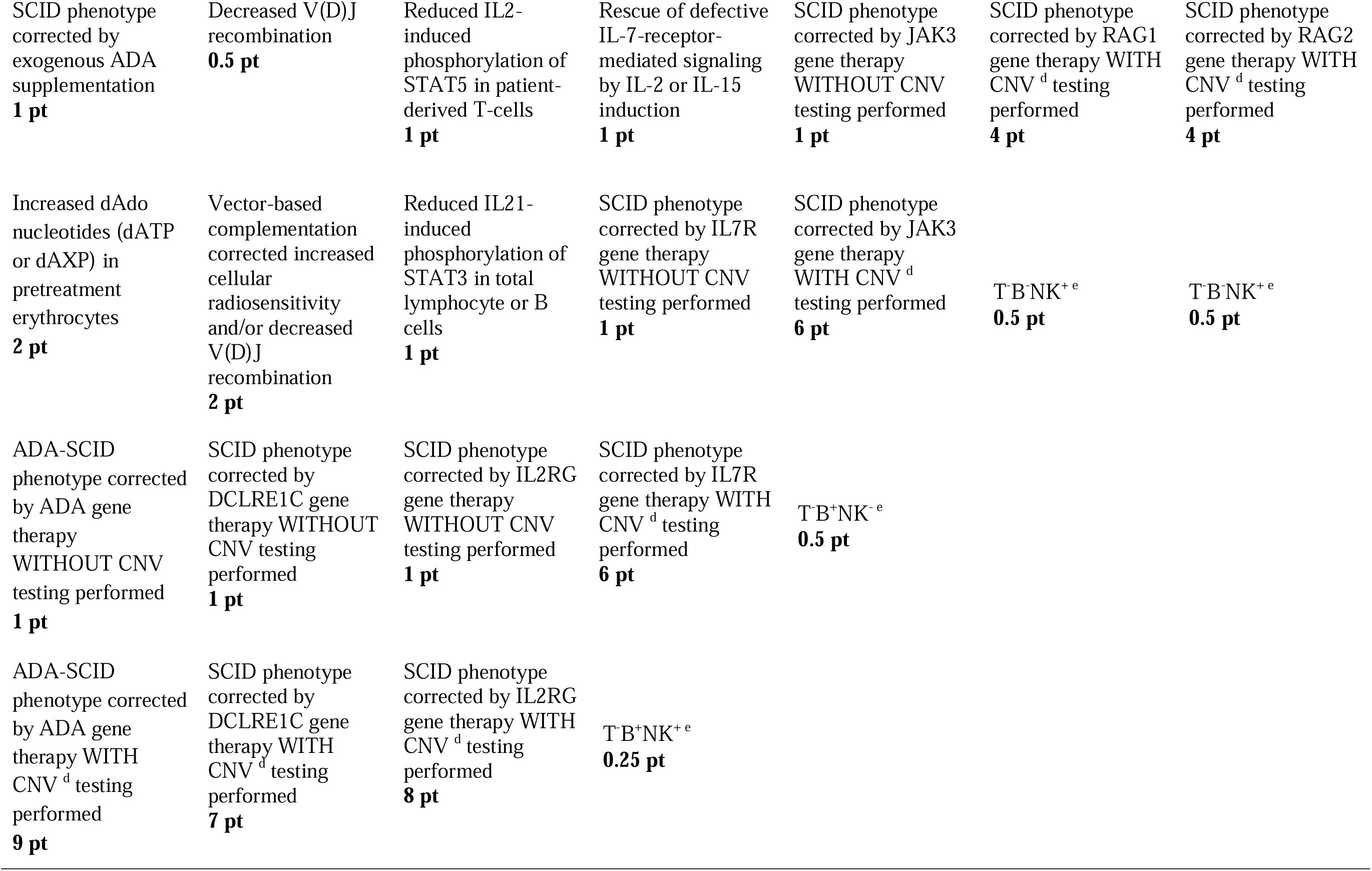
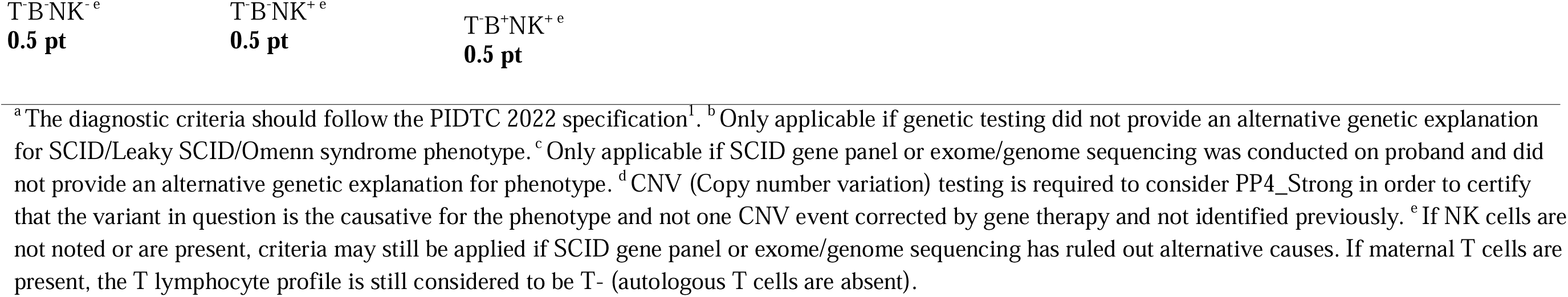
Clinical features deemed highly specific for each gene (PP4)

#### Exclusion of ACMG/AMP criteria

The exclusion criteria that are universally not applicable across all genes studied are collectively presented here: PP5, BP6, BP1, BP2, BP3, BP4, BP5, and PP2.

We adopted the ClinGen advisement against use of clinical laboratory classification (PP5, BP6) ^24^. Furthermore, BP1 (a missense variant in a gene for which primarily truncating variants are known to cause disease) was determined as inapplicable because many pathogenic variants in the 7 genes are indeed missense. This assessment was also based upon ClinVar, where the median percentage of missense variants in the 7 genes was approximately 80%. Additionally, BP2 (observed *in trans* with a pathogenic variant for a fully penetrant dominant gene/disorder or observed *in cis* with a pathogenic variant in any inheritance pattern) was defined as inapplicable. The VCEP felt that possibilities might occur wherein the variant of interest is truly pathogenic but by chance is located *in cis* with a known pathogenic variant along with an unclassified or missing variant on the opposite allele.

Because the frequency or likelihood of such an event is not known, the panel elected to curtail use of BP2.

BP3 (in-frame deletions/insertions in a repetitive region without known function) and BP4 (multiple lines of computational evidence suggest no impact on gene or gene product) were also deemed not applicable, as none of our genes have known repetitive regions and *in silico* prediction data and thresholds have not been validated for the genes.

We also removed BP5 (alternative mechanism for disease), given that in very rare circumstances, a patient can carry defects in two different genes causing SCID.

Finally, the PP2 criterion (missense variant in a gene that has a low rate of benign missense variation and where missense variants are a common mechanism of disease) was also deemed inapplicable. This assertion was based on the low z-score values for all genes (*ADA*=0.46, *DCLRE1C*=0.05, *IL7R*=-1.18, *JAK3*=1.67, *IL2RG*= 1.49, *RAG1*=1.77, and *RAG2*=0.57) in the gnomAD missense constraint table, which fall below the SVI recommendation (>3.09) for applying this criterion, as specified in the ClinGen VCEP Standard Operating Protocol^25^.

## PERFORMANCE OF THE SCID-VCEP ACMG/AMP SPECIFICATIONS IN VARIANT CLASSIFICATION

The modified SCID-VCEP specifications were tested with a pilot set of 90 variants. The variants were selected from ClinVar and from research and private laboratory data. They included variants with previous assertions of benign/likely benign (B/LB), pathogenic/likely pathogenic (P/LP), variant of uncertain significance (VUS), and variants with absent or conflicting interpretations. The selection comprised missense, synonymous, frameshift, splice site, and intronic variants to allow for a comprehensive comparison of how our criteria could be utilized. Case segregation and functional evidence were gathered from limited internal data as well as available published literature.

Utilizing these specifications, 25 variants were classified as pathogenic, 21 as likely pathogenic, 14 as VUS, 18 as likely benign, and 12 as benign. Each of these variants was curated in the ClinGen Variant Curation Interface^13^. The classifications were approved by the general SCID-VCEP and submitted to the ClinGen Evidence Repository and ClinVar for publication. The changes from previous classifications in ClinVar and the SCID-VCEP classifications can be seen in Figure 1. Supplemental Table 4 contains a complete list of all 90 variants curated, as well as all applicable codes (Table S4). Following the pilot phase, we initiated a sustained phase, and as of now, 215 variant classifications have been published in ClinVar.

**Figure 1.**
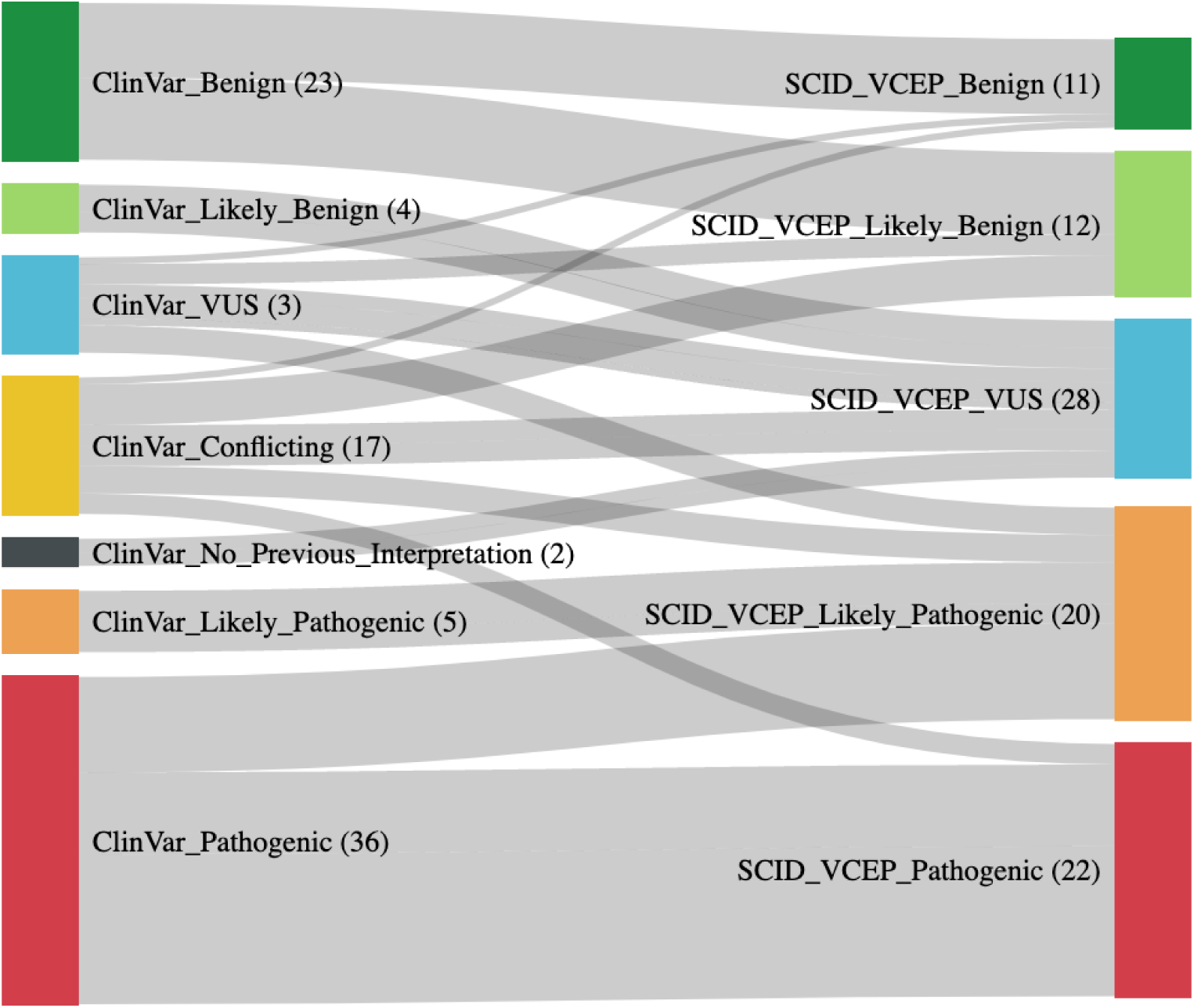
Classification of the 90 selected pilot variants by the original ClinVar assertion (left) and the SCID-VCEP specifics adapted from ACMG/AMP guidelines (right). VUS: Variant of Uncertain Significance. Raw numbers are indicated in parentheses. The variants are approximately equally distributed across the following genes: *ADA* (14), *DCLRE1C* (12), *IL7R* (14), *RAG1* (12), *RAG2* (12), *JAK3* (12), and *IL2RG* (14).

## LIMITATIONS

In evaluating the SCID-VCEP specifications and the variant curation process, it remains essential to recognize the constraints that impact its efficacy and scope. The limitations of this process include the absence of validated thresholds for using *in silico* tools for these genes. This deficiency constrained our ability to apply PP3 and BP7. Moreover, the lack of broad use of established *in vitro* functional assays for specific genes, including *IL2RG*, *IL7R*, and *JAK3*, will limit applicability of the functional codes during most classification attempts. Furthermore, the lack of benign variants tested in publications, prevents application of PS3 functional evidence at a strong level in the ACMG/AMP evidence framework. These challenges underscore the critical need to develop advanced high-throughput functional assays. Such assays would enable a comprehensive analysis of all potential missense variants in these genes. Equally important is the need to validate newer computational algorithms designed to assess DNA alterations and their consequential effects on proteins.

## CONCLUSION AND FUTURE DIRECTIONS

The establishment of the SCID-VCEP represents a significant advancement for the genetic diagnosis of SCID. This initiative is critical for addressing the complex genetic landscape of SCID, where precise variant classification in genes with established relationships to SCID is pivotal for effective patient care.

Application of gene-specific rules for ACMG/AMP variant classification in *ADA*, *DCLRE1C*, *Il2RG*, *IL7R*, *JAK3*, *RAG1*, and *RAG2* has demonstrated encouraging outcomes in a pilot study of 90 variants, with 85% achieving definitive classification (P/LP, B/LB). Regular application of these specifications is likely to decrease the frequency of VUS reported on patients undergoing SCID diagnostic evaluation. Ongoing refinement of these guidelines is anticipated to enhance the clinical utility of variant classification further over time.

The SCID-VCEP variant classification specifications represent a significant step forward in the genetic diagnosis of SCID. By providing a robust framework for variant classification, these specifications enhance the accuracy of genetic testing for patient care. Ongoing refinement and validation of these specifications will ensure that they continue to meet the needs of the clinical community and contribute to better outcomes for patients with SCID.

## Supporting information

PS3 and BS3 Functional Evidence

PVS1 Flowchart

Pilot Phase Results

Supplemental references

## DATA AVAILABILITY

The ClinGen Severe Combined Immunodeficiency Disease Variant Curation Expert Panel submitted all variants to the ClinVar Database^26^ including evidence summaries detailing the data used for each classification. The detailed evidence used for the classification of these variants is available in the ClinGen Evidence Repository^27^. Supplemental Material 3 provides a comprehensive list of variants along with their ClinVar identifiers and the applied codes.

Additionally, it includes a curation summary for all variants.

## MEMBERS OF THE CLINGEN SEVERE COMBINED IMMUNODEFICIENCY VARIANT CURATION EXPERT PANEL

Chairs: Ivan K. Chinn, MD, Britt A. Johnson, PhD, FACMG. Coordinators: Michelle Zelnick, MS, CGC. Curators: Abduarahman Almutairi, MD, Agustin Bernacchia, Anita Chacko, PhD, Mark Dulchavsky, Shirley PY Hue, Neema Izadi, MD, MSc, Vanessa C. Jacovas, PhD, Namrata Khurana, PhD, Grace Leon-Lozano, BS, Wei Ma, Shiloh Martin, MD, PhD, Xueyang Pan, PhD, Justyne E. Ross, PhD, Xinrui Shi, PhD. Members: Eric Allenspach, MD, PhD, Alice Y. Chan, MD, PhD, Brian Chung, MD, PhD, FCCMG, Morton J. Cowan, MD, Rajarshi Ghosh, PhD, FACMG, Mike Hershfield, MD, Xi Luo, PhD, FACMG, Luigi D. Notarangelo, MD, Megan Piazza, PhD, FACMG, Jennifer M. Puck, MD, Katharina V. Schulze, Ph.D., Theru Sivakumaran, PhD, FACMG, Svetlana Vakkilainen, MD, PhD, Nicolai S.C. van Oers, PhD, James Verbsky, MD, PhD, Jolan Walter, MD, PhD, Bo Yuan, PhD, FACMG. Past Members: Jordan Abbott, MD, Marwa Elnagheeb, MPH, Benjamin McLean, BA, Shannon McNulty, PhD, Emma Owens, BS, Brooke Palus, M.S., Jin Peng, MS, Francesco Vetrini, PhD, FACMG.

## CONFLICT OF INTEREST

VCJ: Employee and shareholder of Fulgent Genetics. MZ: No conflict of interest to declare. SM: No conflict of interest to declare. JER: No conflict of interest to declare. NK: No conflict of interest to declare. XP: No conflict of interest to declare. AN: No conflict of interest to declare. SM: Employee and shareholder of Labcorp. BM: No conflict of interest to declare.

MAE: No conflict of interest to declare. MJC: Member of DSMB for Bluebird Bio, Rocket Pharma, and Chiesi Bio; royalties from UpToDate. JMP: Royalties from UpToDate. MH: Grant support from Chiesi USA. JV: No conflict of interest to declare. JW: Orca Bio; Grifols; Takeda; ADMA Biologicals; Octapharma; X4-Pharmaceuticals; Bristol-Myers Squibb.

Consultant/Advisory Boards: Takeda; Pharming; Cheisi; Genpharm Speaker’s Bureau: Pharming; Takeda; Genpharm. Royalties from UptoDate: medical writer. Member of the DSMB (Pharming). EA: No conflict of interest to declare. AYC: Consultant to Sobi. NSCO: No conflict of interest to declare. RG: No conflict of interest to declare. MP: Employee PreventionGenetics, part of Exact Sciences. BY: No conflict of interest to declare. LDN:

Royalties from UpToDate. BAJ: Employee and shareholder of GeneDx, LLC. IKC: Royalties from Wolters Kluwer for UpToDate.

## ACKNOWLEDGMENTS

We thank the Clinical Genome Resource organization for feedback and support during our application. This publication was supported in part by the National Human Genome Research Institute of the National Institutes of Health through U24HG009650. The content is solely the responsibility of the author and does not necessarily represent the official views of the National Institutes of Health. Additionally, this work was supported by the National Institutes of Health (NIH) under a grant from the National Institute of Child Health and Human Development (NICHD) (1U24HD104590-01).

We also extend our gratitude to all the volunteer biocurators and expert panelists.

MJC’s work was supported by California Institute of Regenerative Medicine CLIN2-10830 and NIH U54 AI 082973.

