## Supplementary material for "The ClinGen Severe Combined Immunodeficiency Disease Variant Curation Expert Panel: Specifications for classification of variants in *ADA*, *DCLRE1C*, *IL2RG*, *IL7R*, *JAK3*, *RAG1*, and *RAG2*": PVS1 Flowchart

*ADA:*

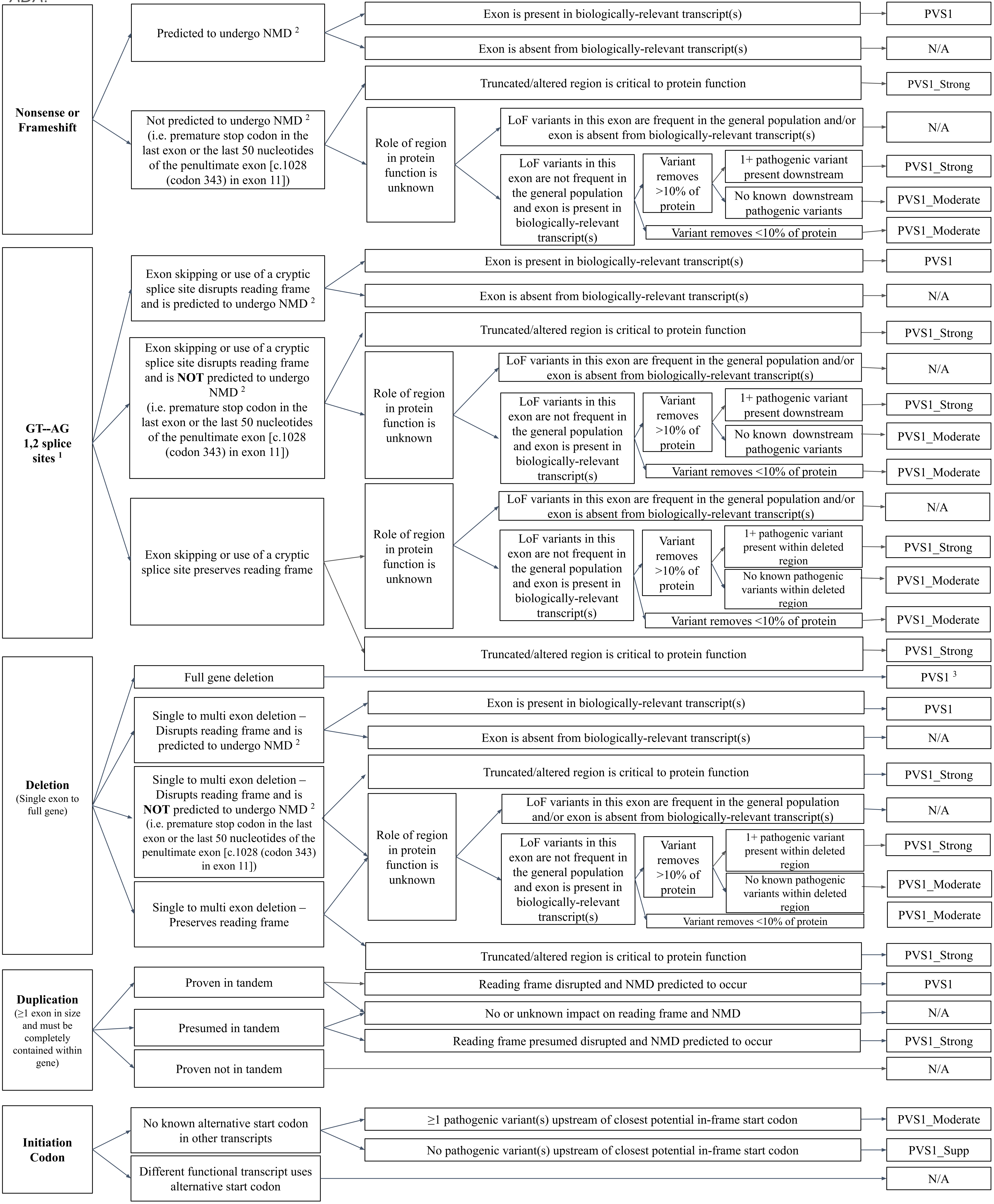

Nonsense or Frameshift
Predicted to undergo NMD 2
Exon is present in biologically-relevant transcript(s)
PVS1
Exon is absent from biologically-relevant transcript(s)
N/A
Not predicted to undergo NMD 2 (i.e. premature stop codon in the last exon or the last 50 nucleotides of the penultimate exon [c.1028 (codon 343) in exon 11])
Truncated/altered region is critical to protein function
PVS1_Strong
Role of region in protein function is unknown
LoF variants in this exon are frequent in the general population and/or exon is absent from biologically-relevant transcript(s)
N/A
LoF variants in this exon are not frequent in the general population and exon is present in biologically-relevant transcript(s)
Variant removes >10% of protein
1+ pathogenic variant present downstream
PVS1_Strong
No known downstream pathogenic variants
PVS1_Moderate
Variant removes <10% of protein
PVS1_Moderate
GT--AG 1,2 splice sites 1
Exon skipping or use of a cryptic splice site disrupts reading frame and is predicted to undergo NMD 2
Exon is present in biologically-relevant transcript(s)
PVS1
Exon is absent from biologically-relevant transcript(s)
N/A
Exon skipping or use of a cryptic splice site disrupts reading frame and is NOT predicted to undergo NMD 2 (i.e. premature stop codon in the last exon or the last 50 nucleotides of the penultimate exon [c.1028 (codon 343) in exon 11])
Truncated/altered region is critical to protein function
PVS1_Strong
Role of region in protein function is unknown
LoF variants in this exon are frequent in the general population and/or exon is absent from biologically-relevant transcript(s)
N/A
LoF variants in this exon are not frequent in the general population and exon is present in biologically-relevant transcript(s)
Variant removes >10% of protein
1+ pathogenic variant present downstream
PVS1_Strong
No known downstream pathogenic variants
PVS1_Moderate
Variant removes <10% of protein
PVS1_Moderate
Exon skipping or use of a cryptic splice site preserves reading frame
Role of region in protein function is unknown
LoF variants in this exon are frequent in the general population and/or exon is absent from biologically-relevant transcript(s)
N/A
LoF variants in this exon are not frequent in the general population and exon is present in biologically-relevant transcript(s)
Variant removes >10% of protein
1+ pathogenic variant present within deleted region
PVS1_Strong
No known pathogenic variants within deleted region
PVS1_Moderate
Variant removes <10% of protein
PVS1_Moderate
Truncated/altered region is critical to protein function
PVS1_Strong
Deletion (Single exon to full gene)
Full gene deletion
PVS1 3
Single to multi exon deletion – Disrupts reading frame and is predicted to undergo NMD 2
Exon is present in biologically-relevant transcript(s)
PVS1
Exon is absent from biologically-relevant transcript(s)
N/A
Single to multi exon deletion – Disrupts reading frame and is NOT predicted to undergo NMD 2 (i.e. premature stop codon in the last exon or the last 50 nucleotides of the penultimate exon [c.1028 (codon 343) in exon 11])
Truncated/altered region is critical to protein function
PVS1_Strong
Role of region in protein function is unknown
LoF variants in this exon are frequent in the general population and/or exon is absent from biologically-relevant transcript(s)
N/A
LoF variants in this exon are not frequent in the general population and exon is present in biologically-relevant transcript(s)
Variant removes >10% of protein
1+ pathogenic variant present within deleted region
PVS1_Strong
No known pathogenic variants within deleted region
PVS1_Moderate
Variant removes <10% of protein
PVS1_Moderate
Single to multi exon deletion – Preserves reading frame
Truncated/altered region is critical to protein function
PVS1_Strong
Duplication (≥1 exon in size and must be completely contained within gene)
Proven in tandem
Presumed in tandem
Proven not in tandem
Reading frame disrupted and NMD predicted to occur
PVS1
No or unknown impact on reading frame and NMD
N/A
Reading frame presumed disrupted and NMD predicted to occur
PVS1_Strong
N/A
Initiation Codon
No known alternative start codon in other transcripts
≥1 pathogenic variant(s) upstream of closest potential in-frame start codon
PVS1_Moderate
No pathogenic variant(s) upstream of closest potential in-frame start codon
PVS1_Supp
Different functional transcript uses alternative start codon
N/A

*DCLRE1C*:

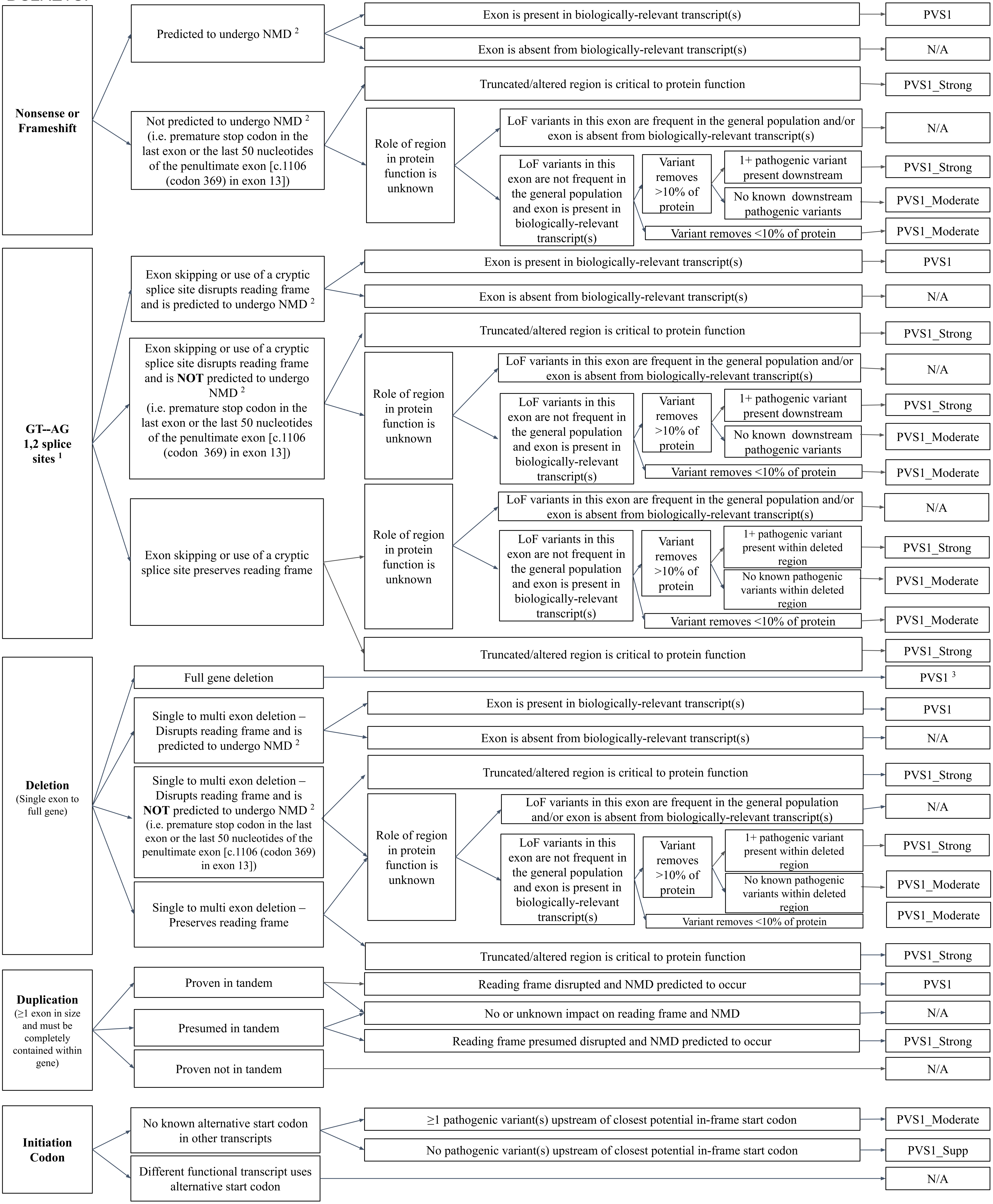

Nonsense or Frameshift
Predicted to undergo NMD [2]
Exon is present in biologically-relevant transcript(s)
PVS1
Exon is absent from biologically-relevant transcript(s)
N/A
Not predicted to undergo NMD [2] (i.e. premature stop codon in the last exon or the last 50 nucleotides of the penultimate exon [c.1106 (codon 369) in exon 13])
Truncated/altered region is critical to protein function
PVS1_Strong
Role of region in protein function is unknown
LoF variants in this exon are frequent in the general population and/or exon is absent from biologically-relevant transcript(s)
N/A
LoF variants in this exon are not frequent in the general population and exon is present in biologically-relevant transcript(s)
Variant removes >10% of protein
1+ pathogenic variant present downstream
PVS1_Strong
No known downstream pathogenic variants
PVS1_Moderate
Variant removes <10% of protein
PVS1_Moderate
GT--AG 1,2 splice sites [1]
Exon skipping or use of a cryptic splice site disrupts reading frame and is predicted to undergo NMD [2]
Exon is present in biologically-relevant transcript(s)
PVS1
Exon is absent from biologically-relevant transcript(s)
N/A
Exon skipping or use of a cryptic splice site disrupts reading frame and is NOT predicted to undergo NMD [2] (i.e. premature stop codon in the last exon or the last 50 nucleotides of the penultimate exon [c.1106 (codon 369) in exon 13])
Truncated/altered region is critical to protein function
PVS1_Strong
Role of region in protein function is unknown
LoF variants in this exon are frequent in the general population and/or exon is absent from biologically-relevant transcript(s)
N/A
LoF variants in this exon are not frequent in the general population and exon is present in biologically-relevant transcript(s)
Variant removes >10% of protein
1+ pathogenic variant present downstream
PVS1_Strong
No known downstream pathogenic variants
PVS1_Moderate
Variant removes <10% of protein
PVS1_Moderate
Exon skipping or use of a cryptic splice site preserves reading frame
Role of region in protein function is unknown
LoF variants in this exon are frequent in the general population and/or exon is absent from biologically-relevant transcript(s)
N/A
LoF variants in this exon are not frequent in the general population and exon is present in biologically-relevant transcript(s)
Variant removes >10% of protein
1+ pathogenic variant present within deleted region
PVS1_Strong
No known pathogenic variants within deleted region
PVS1_Moderate
Variant removes <10% of protein
PVS1_Moderate
Truncated/altered region is critical to protein function
PVS1_Strong
Deletion (Single exon to full gene)
Full gene deletion
PVS1 [3]
Single to multi exon deletion – Disrupts reading frame and is predicted to undergo NMD [2]
Exon is present in biologically-relevant transcript(s)
PVS1
Exon is absent from biologically-relevant transcript(s)
N/A
Single to multi exon deletion – Disrupts reading frame and is NOT predicted to undergo NMD [2] (i.e. premature stop codon in the last exon or the last 50 nucleotides of the penultimate exon [c.1106 (codon 369) in exon 13])
Truncated/altered region is critical to protein function
PVS1_Strong
Role of region in protein function is unknown
LoF variants in this exon are frequent in the general population and/or exon is absent from biologically-relevant transcript(s)
N/A
LoF variants in this exon are not frequent in the general population and exon is present in biologically-relevant transcript(s)
Variant removes >10% of protein
1+ pathogenic variant present within deleted region
PVS1_Strong
No known pathogenic variants within deleted region
PVS1_Moderate
Variant removes <10% of protein
PVS1_Moderate
Single to multi exon deletion – Preserves reading frame
Truncated/altered region is critical to protein function
PVS1_Strong
Duplication (≥1 exon in size and must be completely contained within gene)
Proven in tandem
Reading frame disrupted and NMD predicted to occur
PVS1
Presumed in tandem
No or unknown impact on reading frame and NMD
N/A
Reading frame presumed disrupted and NMD predicted to occur
PVS1_Strong
Proven not in tandem
N/A
Initiation Codon
No known alternative start codon in other transcripts
≥1 pathogenic variant(s) upstream of closest potential in-frame start codon
PVS1_Moderate
No pathogenic variant(s) upstream of closest potential in-frame start codon
PVS1_Supp
Different functional transcript uses alternative start codon
N/A

*IL2RG*:

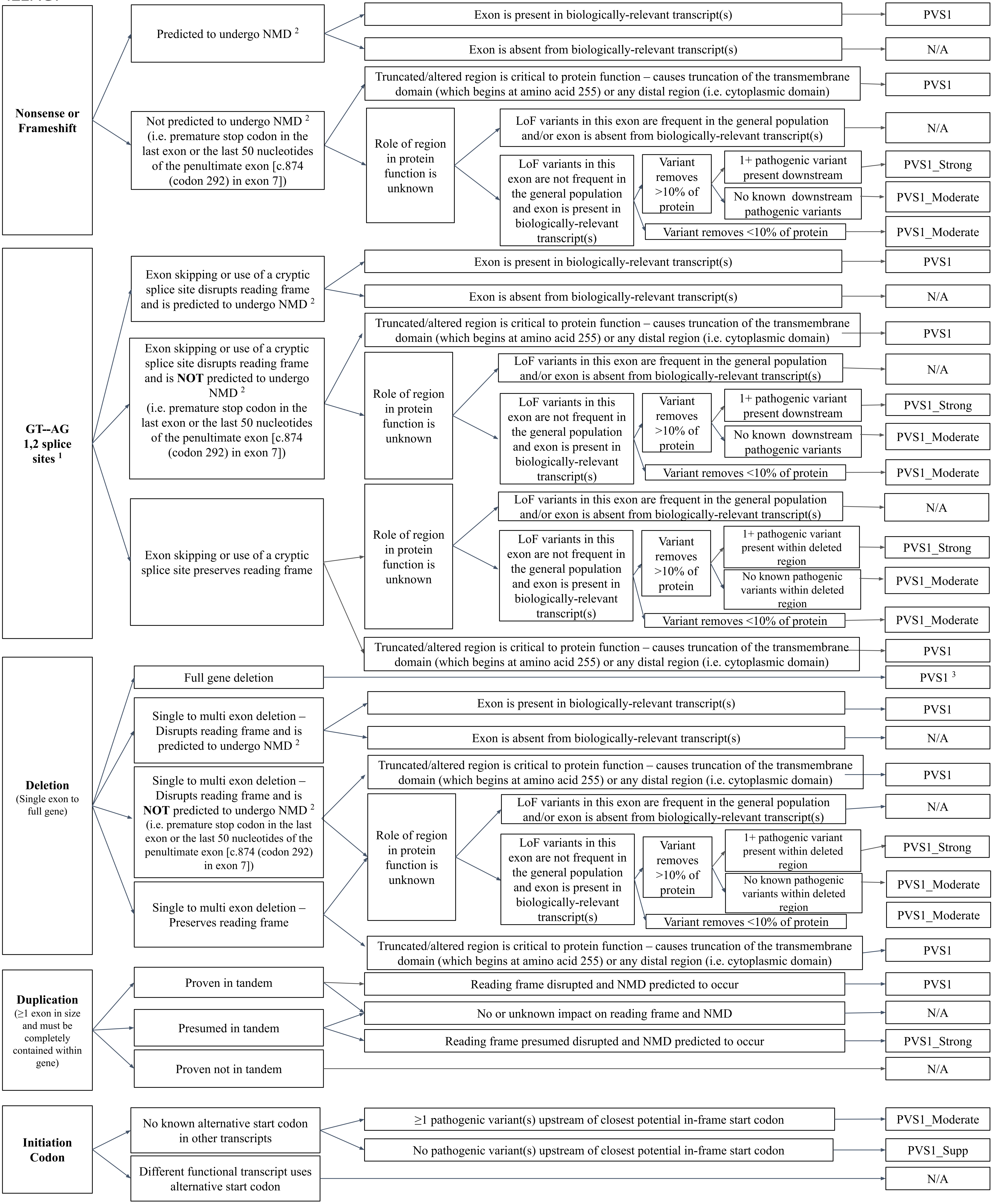

Nonsense or Frameshift
Predicted to undergo NMD [2]
Exon is present in biologically-relevant transcript(s)
PVS1
Exon is absent from biologically-relevant transcript(s)
N/A
Not predicted to undergo NMD [2] (i.e. premature stop codon in the last exon or the last 50 nucleotides of the penultimate exon [c.874 (codon 292) in exon 7])
Truncated/altered region is critical to protein function – causes truncation of the transmembrane domain (which begins at amino acid 255) or any distal region (i.e. cytoplasmic domain)
PVS1
Role of region in protein function is unknown
LoF variants in this exon are frequent in the general population and/or exon is absent from biologically-relevant transcript(s)
N/A
LoF variants in this exon are not frequent in the general population and exon is present in biologically-relevant transcript(s)
Variant removes >10% of protein
1+ pathogenic variant present downstream
PVS1_Strong
No known downstream pathogenic variants
PVS1_Moderate
Variant removes <10% of protein
PVS1_Moderate
GT--AG 1,2 splice sites [1]
Exon skipping or use of a cryptic splice site disrupts reading frame and is predicted to undergo NMD [2]
Exon is present in biologically-relevant transcript(s)
PVS1
Exon is absent from biologically-relevant transcript(s)
N/A
Exon skipping or use of a cryptic splice site disrupts reading frame and is NOT predicted to undergo NMD [2] (i.e. premature stop codon in the last exon or the last 50 nucleotides of the penultimate exon [c.874 (codon 292) in exon 7])
Truncated/altered region is critical to protein function – causes truncation of the transmembrane domain (which begins at amino acid 255) or any distal region (i.e. cytoplasmic domain)
PVS1
Role of region in protein function is unknown
LoF variants in this exon are frequent in the general population and/or exon is absent from biologically-relevant transcript(s)
N/A
LoF variants in this exon are not frequent in the general population and exon is present in biologically-relevant transcript(s)
Variant removes >10% of protein
1+ pathogenic variant present downstream
PVS1_Strong
No known downstream pathogenic variants
PVS1_Moderate
Variant removes <10% of protein
PVS1_Moderate
Exon skipping or use of a cryptic splice site preserves reading frame
Role of region in protein function is unknown
LoF variants in this exon are frequent in the general population and/or exon is absent from biologically-relevant transcript(s)
N/A
LoF variants in this exon are not frequent in the general population and exon is present in biologically-relevant transcript(s)
Variant removes >10% of protein
1+ pathogenic variant present within deleted region
PVS1_Strong
No known pathogenic variants within deleted region
PVS1_Moderate
Variant removes <10% of protein
PVS1_Moderate
Truncated/altered region is critical to protein function – causes truncation of the transmembrane domain (which begins at amino acid 255) or any distal region (i.e. cytoplasmic domain)
PVS1
Deletion (Single exon to full gene)
Full gene deletion
PVS1 [3]
Single to multi exon deletion – Disrupts reading frame and is predicted to undergo NMD [2]
Exon is present in biologically-relevant transcript(s)
PVS1
Exon is absent from biologically-relevant transcript(s)
N/A
Single to multi exon deletion – Disrupts reading frame and is NOT predicted to undergo NMD [2] (i.e. premature stop codon in the last exon or the last 50 nucleotides of the penultimate exon [c.874 (codon 292) in exon 7])
Truncated/altered region is critical to protein function – causes truncation of the transmembrane domain (which begins at amino acid 255) or any distal region (i.e. cytoplasmic domain)
PVS1
Role of region in protein function is unknown
LoF variants in this exon are frequent in the general population and/or exon is absent from biologically-relevant transcript(s)
N/A
LoF variants in this exon are not frequent in the general population and exon is present in biologically-relevant transcript(s)
Variant removes >10% of protein
1+ pathogenic variant present within deleted region
PVS1_Strong
No known pathogenic variants within deleted region
PVS1_Moderate
Variant removes <10% of protein
PVS1_Moderate
Single to multi exon deletion – Preserves reading frame
Truncated/altered region is critical to protein function – causes truncation of the transmembrane domain (which begins at amino acid 255) or any distal region (i.e. cytoplasmic domain)
PVS1
Duplication (≥1 exon in size and must be completely contained within gene)
Proven in tandem
Presumed in tandem
Proven not in tandem
Reading frame disrupted and NMD predicted to occur
PVS1
No or unknown impact on reading frame and NMD
N/A
Reading frame presumed disrupted and NMD predicted to occur
PVS1_Strong
N/A
Initiation Codon
No known alternative start codon in other transcripts
≥1 pathogenic variant(s) upstream of closest potential in-frame start codon
PVS1_Moderate
No pathogenic variant(s) upstream of closest potential in-frame start codon
PVS1_Supp
Different functional transcript uses alternative start codon
N/A

*IL7R*:

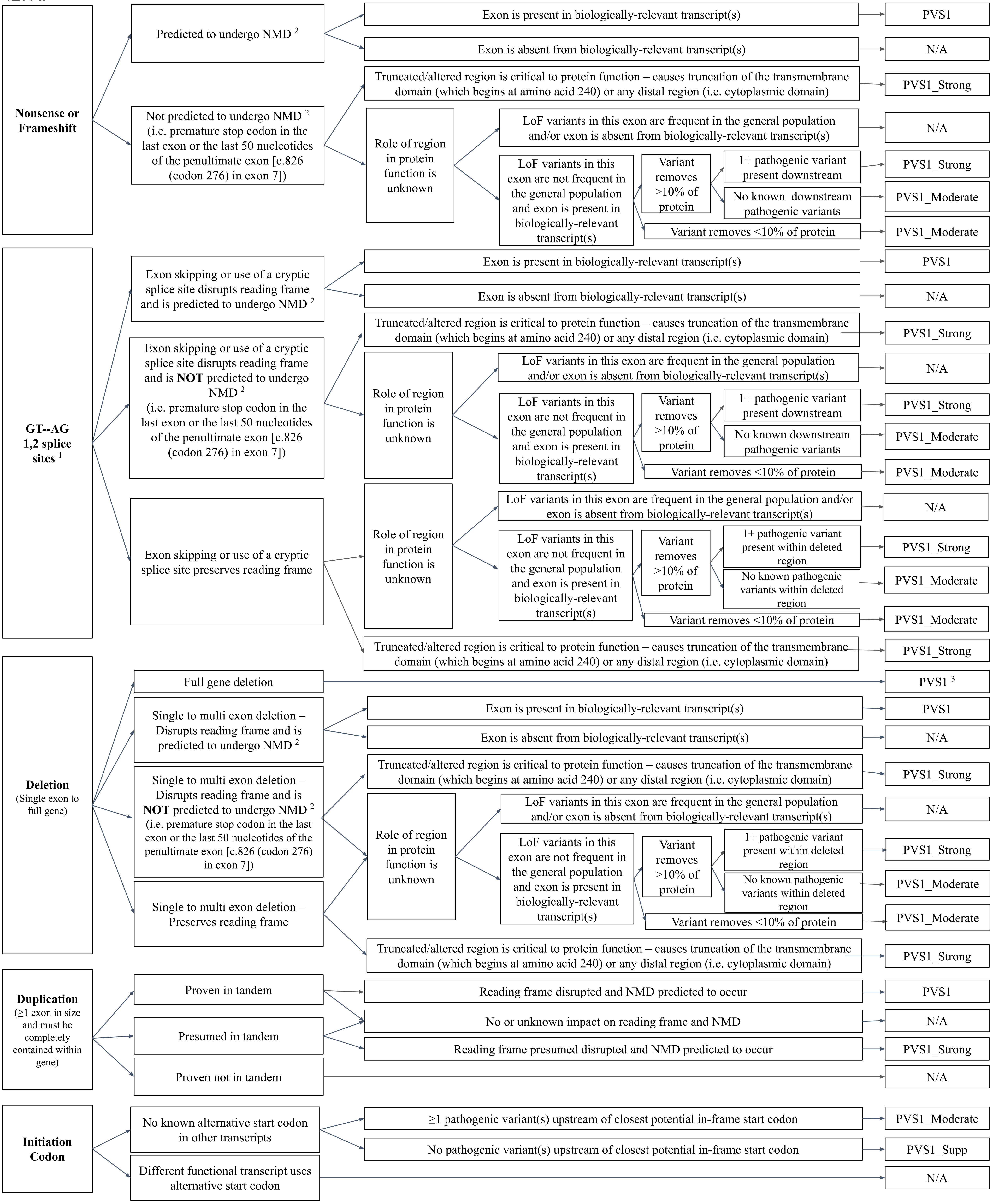
Nonsense or Frameshift
Predicted to undergo NMD [2]
Exon is present in biologically-relevant transcript(s)
PVS1
Exon is absent from biologically-relevant transcript(s)
N/A
Not predicted to undergo NMD [2] (i.e. premature stop codon in the last exon or the last 50 nucleotides of the penultimate exon [c.826 (codon 276) in exon 7])
Truncated/altered region is critical to protein function – causes truncation of the transmembrane domain (which begins at amino acid 240) or any distal region (i.e. cytoplasmic domain)
PVS1_Strong
Role of region in protein function is unknown
LoF variants in this exon are frequent in the general population and/or exon is absent from biologically-relevant transcript(s)
N/A
LoF variants in this exon are not frequent in the general population and exon is present in biologically-relevant transcript(s)
Variant removes >10% of protein
1+ pathogenic variant present downstream
PVS1_Strong
No known downstream pathogenic variants
PVS1_Moderate
Variant removes <10% of protein
PVS1_Moderate
GT--AG 1,2 splice sites [1]
Exon skipping or use of a cryptic splice site disrupts reading frame and is predicted to undergo NMD [2]
Exon is present in biologically-relevant transcript(s)
PVS1
Exon is absent from biologically-relevant transcript(s)
N/A
Exon skipping or use of a cryptic splice site disrupts reading frame and is NOT predicted to undergo NMD [2] (i.e. premature stop codon in the last exon or the last 50 nucleotides of the penultimate exon [c.826 (codon 276) in exon 7])
Truncated/altered region is critical to protein function – causes truncation of the transmembrane domain (which begins at amino acid 240) or any distal region (i.e. cytoplasmic domain)
PVS1_Strong
Role of region in protein function is unknown
LoF variants in this exon are frequent in the general population and/or exon is absent from biologically-relevant transcript(s)
N/A
LoF variants in this exon are not frequent in the general population and exon is present in biologically-relevant transcript(s)
Variant removes >10% of protein
1+ pathogenic variant present downstream
PVS1_Strong
No known downstream pathogenic variants
PVS1_Moderate
Variant removes <10% of protein
PVS1_Moderate
Exon skipping or use of a cryptic splice site preserves reading frame
Role of region in protein function is unknown
LoF variants in this exon are frequent in the general population and/or exon is absent from biologically-relevant transcript(s)
N/A
LoF variants in this exon are not frequent in the general population and exon is present in biologically-relevant transcript(s)
Variant removes >10% of protein
1+ pathogenic variant present within deleted region
PVS1_Strong
No known pathogenic variants within deleted region
PVS1_Moderate
Variant removes <10% of protein
PVS1_Moderate
Truncated/altered region is critical to protein function – causes truncation of the transmembrane domain (which begins at amino acid 240) or any distal region (i.e. cytoplasmic domain)
PVS1_Strong
Deletion (Single exon to full gene)
Full gene deletion
PVS1 [3]
Single to multi exon deletion – Disrupts reading frame and is predicted to undergo NMD [2]
Exon is present in biologically-relevant transcript(s)
PVS1
Exon is absent from biologically-relevant transcript(s)
N/A
Single to multi exon deletion – Disrupts reading frame and is NOT predicted to undergo NMD [2] (i.e. premature stop codon in the last exon or the last 50 nucleotides of the penultimate exon [c.826 (codon 276) in exon 7])
Truncated/altered region is critical to protein function – causes truncation of the transmembrane domain (which begins at amino acid 240) or any distal region (i.e. cytoplasmic domain)
PVS1_Strong
Role of region in protein function is unknown
LoF variants in this exon are frequent in the general population and/or exon is absent from biologically-relevant transcript(s)
N/A
LoF variants in this exon are not frequent in the general population and exon is present in biologically-relevant transcript(s)
Variant removes >10% of protein
1+ pathogenic variant present within deleted region
PVS1_Strong
No known pathogenic variants within deleted region
PVS1_Moderate
Variant removes <10% of protein
PVS1_Moderate
Single to multi exon deletion – Preserves reading frame
Truncated/altered region is critical to protein function – causes truncation of the transmembrane domain (which begins at amino acid 240) or any distal region (i.e. cytoplasmic domain)
PVS1_Strong
Duplication (≥1 exon in size and must be completely contained within gene)
Proven in tandem
Presumed in tandem
Proven not in tandem
Reading frame disrupted and NMD predicted to occur
PVS1
No or unknown impact on reading frame and NMD
N/A
Reading frame presumed disrupted and NMD predicted to occur
PVS1_Strong
N/A
Initiation Codon
No known alternative start codon in other transcripts
≥1 pathogenic variant(s) upstream of closest potential in-frame start codon
PVS1_Moderate
No pathogenic variant(s) upstream of closest potential in-frame start codon
PVS1_Supp
Different functional transcript uses alternative start codon
N/A

*JAK3*:

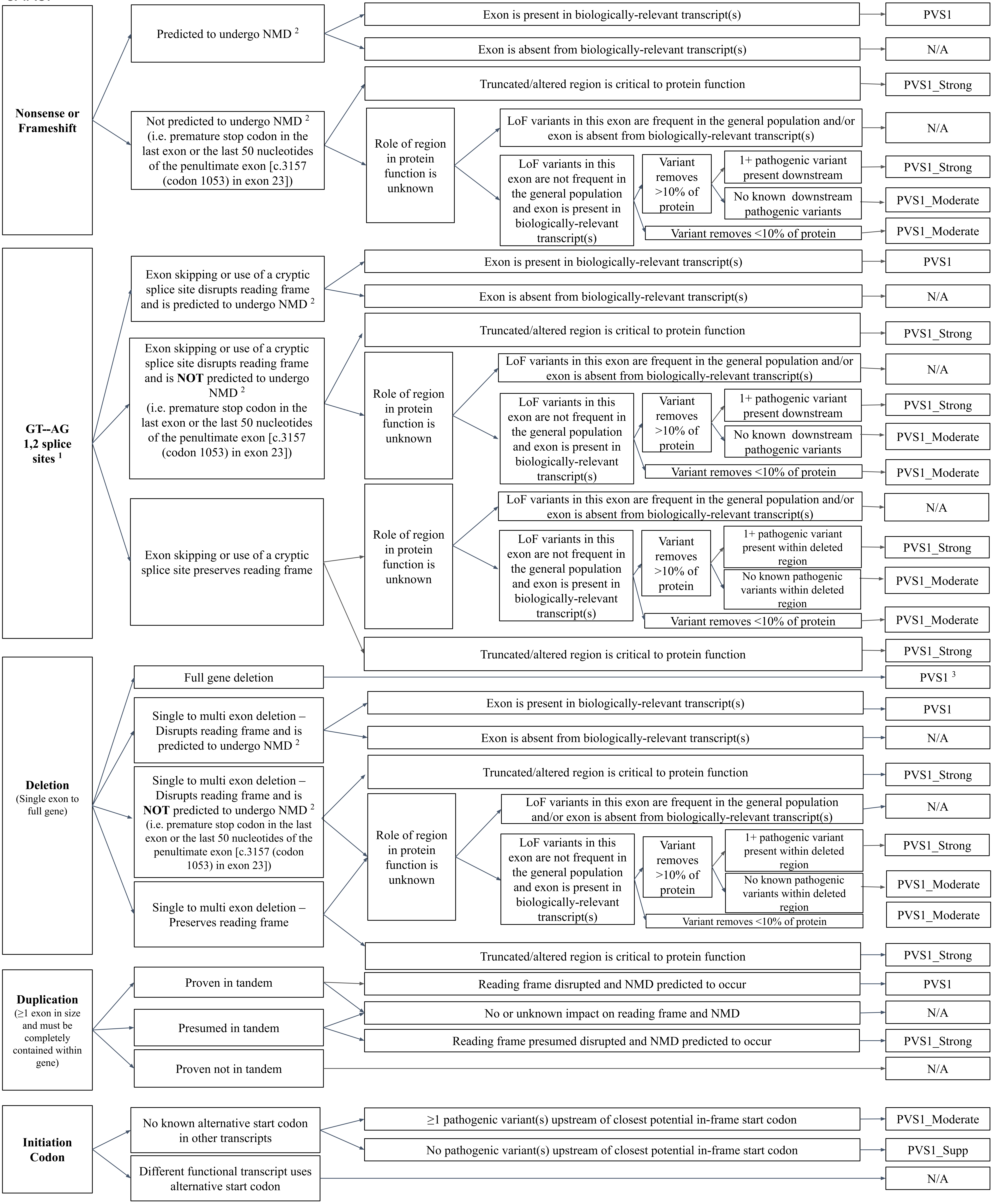
Nonsense or Frameshift
Predicted to undergo NMD [2]
Exon is present in biologically-relevant transcript(s)
PVS1
Exon is absent from biologically-relevant transcript(s)
N/A
Not predicted to undergo NMD [2] (i.e. premature stop codon in the last exon or the last 50 nucleotides of the penultimate exon [c.3157 (codon 1053) in exon 23])
Truncated/altered region is critical to protein function
PVS1_Strong
Role of region in protein function is unknown
LoF variants in this exon are frequent in the general population and/or exon is absent from biologically-relevant transcript(s)
N/A
LoF variants in this exon are not frequent in the general population and exon is present in biologically-relevant transcript(s)
Variant removes >10% of protein
1+ pathogenic variant present downstream
PVS1_Strong
No known downstream pathogenic variants
PVS1_Moderate
Variant removes <10% of protein
PVS1_Moderate
GT--AG 1,2 splice sites [1]
Exon skipping or use of a cryptic splice site disrupts reading frame and is predicted to undergo NMD [2]
Exon is present in biologically-relevant transcript(s)
PVS1
Exon is absent from biologically-relevant transcript(s)
N/A
Exon skipping or use of a cryptic splice site disrupts reading frame and is NOT predicted to undergo NMD [2] (i.e. premature stop codon in the last exon or the last 50 nucleotides of the penultimate exon [c.3157 (codon 1053) in exon 23])
Truncated/altered region is critical to protein function
PVS1_Strong
Role of region in protein function is unknown
LoF variants in this exon are frequent in the general population and/or exon is absent from biologically-relevant transcript(s)
N/A
LoF variants in this exon are not frequent in the general population and exon is present in biologically-relevant transcript(s)
Variant removes >10% of protein
1+ pathogenic variant present downstream
PVS1_Strong
No known downstream pathogenic variants
PVS1_Moderate
Variant removes <10% of protein
PVS1_Moderate
Exon skipping or use of a cryptic splice site preserves reading frame
Role of region in protein function is unknown
LoF variants in this exon are frequent in the general population and/or exon is absent from biologically-relevant transcript(s)
N/A
LoF variants in this exon are not frequent in the general population and exon is present in biologically-relevant transcript(s)
Variant removes >10% of protein
1+ pathogenic variant present within deleted region
PVS1_Strong
No known pathogenic variants within deleted region
PVS1_Moderate
Variant removes <10% of protein
PVS1_Moderate
Truncated/altered region is critical to protein function
PVS1_Strong
Deletion (Single exon to full gene)
Full gene deletion
PVS1 [3]
Single to multi exon deletion – Disrupts reading frame and is predicted to undergo NMD [2]
Exon is present in biologically-relevant transcript(s)
PVS1
Exon is absent from biologically-relevant transcript(s)
N/A
Single to multi exon deletion – Disrupts reading frame and is NOT predicted to undergo NMD [2] (i.e. premature stop codon in the last exon or the last 50 nucleotides of the penultimate exon [c.3157 (codon 1053) in exon 23])
Truncated/altered region is critical to protein function
PVS1_Strong
Role of region in protein function is unknown
LoF variants in this exon are frequent in the general population and/or exon is absent from biologically-relevant transcript(s)
N/A
LoF variants in this exon are not frequent in the general population and exon is present in biologically-relevant transcript(s)
Variant removes >10% of protein
1+ pathogenic variant present within deleted region
PVS1_Strong
No known pathogenic variants within deleted region
PVS1_Moderate
Variant removes <10% of protein
PVS1_Moderate
Single to multi exon deletion – Preserves reading frame
Truncated/altered region is critical to protein function
PVS1_Strong
Duplication (≥1 exon in size and must be completely contained within gene)
Proven in tandem
Reading frame disrupted and NMD predicted to occur
PVS1
Presumed in tandem
No or unknown impact on reading frame and NMD
N/A
Reading frame presumed disrupted and NMD predicted to occur
PVS1_Strong
Proven not in tandem
N/A
Initiation Codon
No known alternative start codon in other transcripts
≥1 pathogenic variant(s) upstream of closest potential in-frame start codon
PVS1_Moderate
No pathogenic variant(s) upstream of closest potential in-frame start codon
PVS1_Supp
Different functional transcript uses alternative start codon
N/A

*RAG1:*

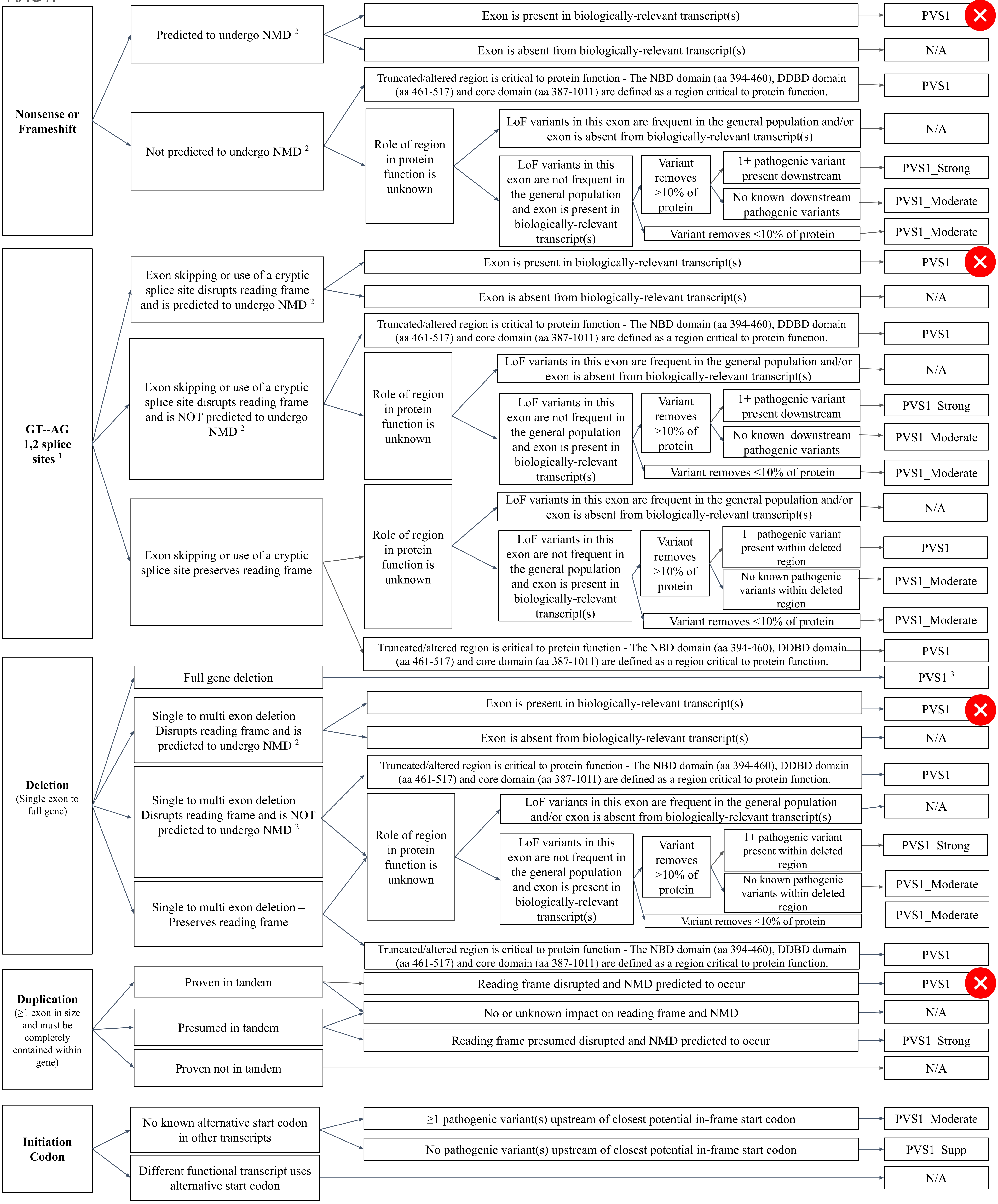

Nonsense or Frameshift
Predicted to undergo NMD 2
Exon is present in biologically-relevant transcript(s)
PVS1
Exon is absent from biologically-relevant transcript(s)
N/A
Not predicted to undergo NMD 2
Truncated/altered region is critical to protein function - The NBD domain (aa 394-460), DDBD domain (aa 461-517) and core domain (aa 387-1011) are defined as a region critical to protein function.
PVS1
Role of region in protein function is unknown
LoF variants in this exon are frequent in the general population and/or exon is absent from biologically-relevant transcript(s)
N/A
LoF variants in this exon are not frequent in the general population and exon is present in biologically-relevant transcript(s)
Variant removes >10% of protein
1+ pathogenic variant present downstream
PVS1_Strong
No known downstream pathogenic variants
PVS1_Moderate
Variant removes <10% of protein
PVS1_Moderate
GT--AG 1,2 splice sites 1
Exon skipping or use of a cryptic splice site disrupts reading frame and is predicted to undergo NMD 2
Exon is present in biologically-relevant transcript(s)
PVS1
Exon is absent from biologically-relevant transcript(s)
N/A
Exon skipping or use of a cryptic splice site disrupts reading frame and is NOT predicted to undergo NMD 2
Truncated/altered region is critical to protein function - The NBD domain (aa 394-460), DDBD domain (aa 461-517) and core domain (aa 387-1011) are defined as a region critical to protein function.
PVS1
Role of region in protein function is unknown
LoF variants in this exon are frequent in the general population and/or exon is absent from biologically-relevant transcript(s)
N/A
LoF variants in this exon are not frequent in the general population and exon is present in biologically-relevant transcript(s)
Variant removes >10% of protein
1+ pathogenic variant present downstream
PVS1_Strong
No known downstream pathogenic variants
PVS1_Moderate
Variant removes <10% of protein
PVS1_Moderate
Exon skipping or use of a cryptic splice site preserves reading frame
Role of region in protein function is unknown
LoF variants in this exon are frequent in the general population and/or exon is absent from biologically-relevant transcript(s)
N/A
LoF variants in this exon are not frequent in the general population and exon is present in biologically-relevant transcript(s)
Variant removes >10% of protein
1+ pathogenic variant present within deleted region
PVS1
No known pathogenic variants within deleted region
PVS1_Moderate
Variant removes <10% of protein
PVS1_Moderate
Truncated/altered region is critical to protein function - The NBD domain (aa 394-460), DDBD domain (aa 461-517) and core domain (aa 387-1011) are defined as a region critical to protein function.
PVS1
Deletion (Single exon to full gene)
Full gene deletion
PVS1 3
Single to multi exon deletion – Disrupts reading frame and is predicted to undergo NMD 2
Exon is present in biologically-relevant transcript(s)
PVS1
Exon is absent from biologically-relevant transcript(s)
N/A
Single to multi exon deletion – Disrupts reading frame and is NOT predicted to undergo NMD 2
Truncated/altered region is critical to protein function - The NBD domain (aa 394-460), DDBD domain (aa 461-517) and core domain (aa 387-1011) are defined as a region critical to protein function.
PVS1
Role of region in protein function is unknown
LoF variants in this exon are frequent in the general population and/or exon is absent from biologically-relevant transcript(s)
N/A
LoF variants in this exon are not frequent in the general population and exon is present in biologically-relevant transcript(s)
Variant removes >10% of protein
1+ pathogenic variant present within deleted region
PVS1_Strong
No known pathogenic variants within deleted region
PVS1_Moderate
Variant removes <10% of protein
PVS1_Moderate
Single to multi exon deletion – Preserves reading frame
Truncated/altered region is critical to protein function - The NBD domain (aa 394-460), DDBD domain (aa 461-517) and core domain (aa 387-1011) are defined as a region critical to protein function.
PVS1
Duplication (≥1 exon in size and must be completely contained within gene)
Proven in tandem
Presumed in tandem
Proven not in tandem
Reading frame disrupted and NMD predicted to occur
PVS1
No or unknown impact on reading frame and NMD
N/A
Reading frame presumed disrupted and NMD predicted to occur
PVS1_Strong
N/A
Initiation Codon
No known alternative start codon in other transcripts
≥1 pathogenic variant(s) upstream of closest potential in-frame start codon
PVS1_Moderate
No pathogenic variant(s) upstream of closest potential in-frame start codon
PVS1_Supp
Different functional transcript uses alternative start codon
N/A

*RAG2:*

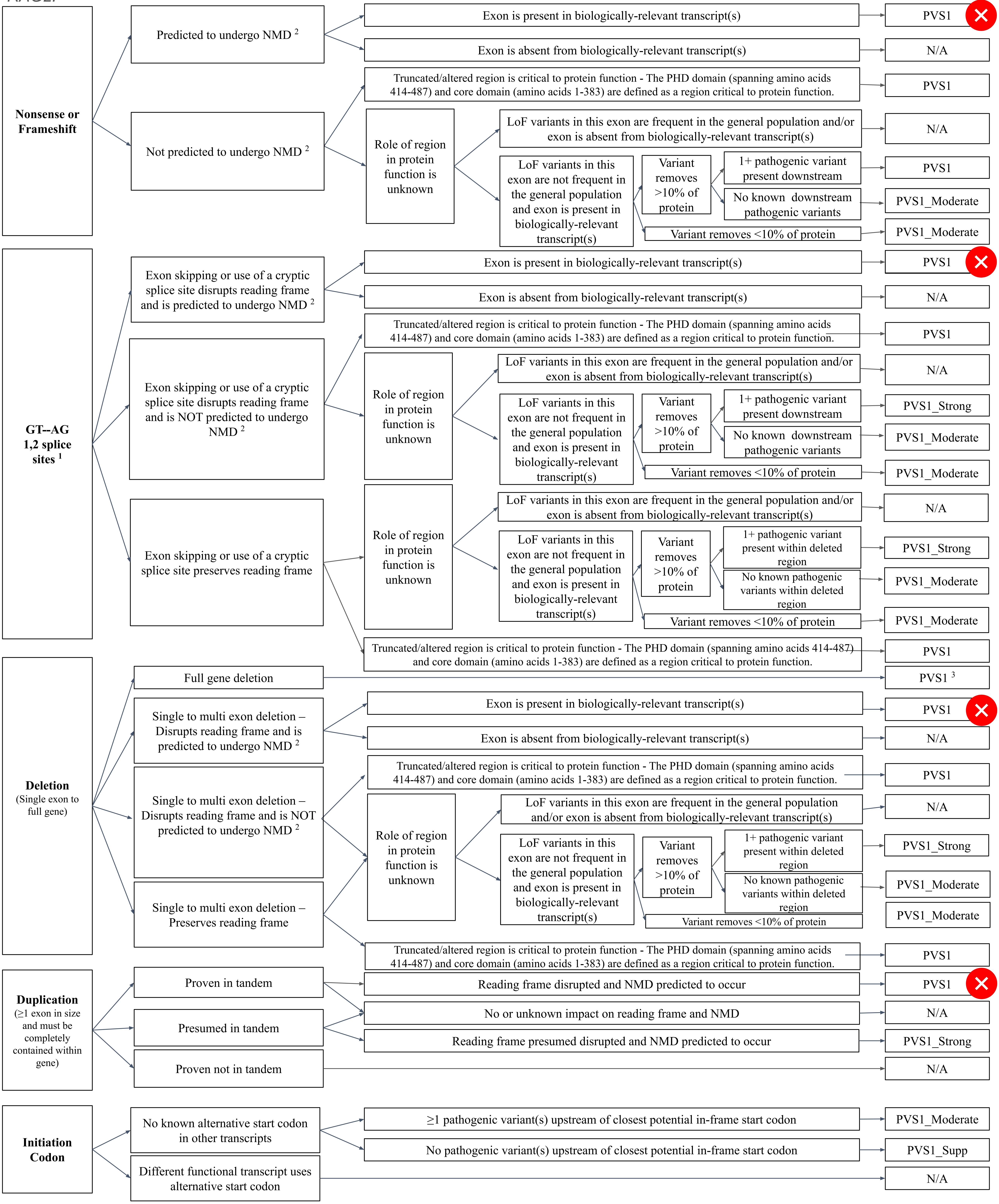

Nonsense or Frameshift
Predicted to undergo NMD 2
Exon is present in biologically-relevant transcript(s)
PVS1
Exon is absent from biologically-relevant transcript(s)
N/A
Not predicted to undergo NMD 2
Truncated/altered region is critical to protein function - The PHD domain (spanning amino acids 414-487) and core domain (amino acids 1-383) are defined as a region critical to protein function.
PVS1
Role of region in protein function is unknown
LoF variants in this exon are frequent in the general population and/or exon is absent from biologically-relevant transcript(s)
N/A
LoF variants in this exon are not frequent in the general population and exon is present in biologically-relevant transcript(s)
Variant removes >10% of protein
1+ pathogenic variant present downstream
PVS1
No known downstream pathogenic variants
PVS1_Moderate
Variant removes <10% of protein
PVS1_Moderate
GT--AG 1,2 splice sites 1
Exon skipping or use of a cryptic splice site disrupts reading frame and is predicted to undergo NMD 2
Exon is present in biologically-relevant transcript(s)
PVS1
Exon is absent from biologically-relevant transcript(s)
N/A
Exon skipping or use of a cryptic splice site disrupts reading frame and is NOT predicted to undergo NMD 2
Truncated/altered region is critical to protein function - The PHD domain (spanning amino acids 414-487) and core domain (amino acids 1-383) are defined as a region critical to protein function.
PVS1
Role of region in protein function is unknown
LoF variants in this exon are frequent in the general population and/or exon is absent from biologically-relevant transcript(s)
N/A
LoF variants in this exon are not frequent in the general population and exon is present in biologically-relevant transcript(s)
Variant removes >10% of protein
1+ pathogenic variant present downstream
PVS1_Strong
No known downstream pathogenic variants
PVS1_Moderate
Variant removes <10% of protein
PVS1_Moderate
Exon skipping or use of a cryptic splice site preserves reading frame
Role of region in protein function is unknown
LoF variants in this exon are frequent in the general population and/or exon is absent from biologically-relevant transcript(s)
N/A
LoF variants in this exon are not frequent in the general population and exon is present in biologically-relevant transcript(s)
Variant removes >10% of protein
1+ pathogenic variant present within deleted region
PVS1_Strong
No known pathogenic variants within deleted region
PVS1_Moderate
Variant removes <10% of protein
PVS1_Moderate
Truncated/altered region is critical to protein function - The PHD domain (spanning amino acids 414-487) and core domain (amino acids 1-383) are defined as a region critical to protein function.
PVS1
Deletion
(Single exon to full gene)
Full gene deletion
PVS1 3
Single to multi exon deletion – Disrupts reading frame and is predicted to undergo NMD 2
Exon is present in biologically-relevant transcript(s)
PVS1
Exon is absent from biologically-relevant transcript(s)
N/A
Single to multi exon deletion – Disrupts reading frame and is NOT predicted to undergo NMD 2
Truncated/altered region is critical to protein function - The PHD domain (spanning amino acids 414-487) and core domain (amino acids 1-383) are defined as a region critical to protein function.
PVS1
Role of region in protein function is unknown
LoF variants in this exon are frequent in the general population and/or exon is absent from biologically-relevant transcript(s)
N/A
LoF variants in this exon are not frequent in the general population and exon is present in biologically-relevant transcript(s)
Variant removes >10% of protein
1+ pathogenic variant present within deleted region
PVS1_Strong
No known pathogenic variants within deleted region
PVS1_Moderate
Variant removes <10% of protein
PVS1_Moderate
Single to multi exon deletion – Preserves reading frame
Truncated/altered region is critical to protein function - The PHD domain (spanning amino acids 414-487) and core domain (amino acids 1-383) are defined as a region critical to protein function.
PVS1
Duplication
(≥1 exon in size and must be completely contained within gene)
Proven in tandem
Reading frame disrupted and NMD predicted to occur
PVS1
Presumed in tandem
No or unknown impact on reading frame and NMD
N/A
Reading frame presumed disrupted and NMD predicted to occur
PVS1_Strong
Proven not in tandem
N/A
Initiation Codon
No known alternative start codon in other transcripts
≥1 pathogenic variant(s) upstream of closest potential in-frame start codon
PVS1_Moderate
No pathogenic variant(s) upstream of closest potential in-frame start codon
PVS1_Supp
Different functional transcript uses alternative start codon
N/A

NMD, nonsense-mediated decay; LoF, loss of function.

1, This criterion should not be applied in combination with in silico splicing predictions (PP3). Additionally, splice site variants must have no detectable nearby (+/− 20 nucleotides) strong consensus splice sequence that may reconstitute in-frame splicing.
2, NMD prediction based on the premature termination codon not occurring in the 3' most exon or the 3'-most 50 bp of the penultimate exon.
3, For a full gene deletion of a known haploinsufficient gene, a Pathogenic classification is warranted (in the absence of conflicting data) even though application of PVS1 alone would not reach a Pathogenic classification using the combining rules in Richards et al 2017.
