## Supplemental tables for "The ClinGen Severe Combined Immunodeficiency Disease Variant Curation Expert Panel: Specifications for classification of variants in *ADA*, *DCLRE1C*, *IL2RG*, *IL7R*, *JAK3*, *RAG1*, and *RAG2*"

Supplemental Table S1. Adapted from “The diagnostic of Severe Combined Immunodeficiency (SCID): The Primary Immune Deficiency Treatment Consortium 2022 Definitions^19^”:

| **SCID subtype** | **Diagnosis requires** | **Criterion 1** | **Criterion 2** | **Criterion 3** | **Criterion 4** |
| --- | --- | --- | --- | --- | --- |
| **Typical SCID**  (very low autologous T cells) | Criteria 1 & 2  **OR** Criteria 1 & 3  **OR** Criterion 4 | Very low T cells (<0.05 × 10^9^/L = <50μL)***** | At least a likely pathogenic variant[**†**](https://www.jacionline.org/article/S0091-6749(22)01479-8/fulltext) | No alternate explanation for low T-cell count[**‡**](https://www.jacionline.org/article/S0091-6749(22)01479-8/fulltext)  **AND, EITHER:**  Undetectable or low TRECs[**§**](https://www.jacionline.org/article/S0091-6749(22)01479-8/fulltext)  **OR** <20% of CD4+ T cells have naive cell surface markers[**\|\|**](https://www.sciencedirect.com/science/article/pii/S0091674922014798#tbl1fnparpar) | Presence of TME[**¶**](https://www.jacionline.org/article/S0091-6749(22)01479-8/fulltext)  (Transplacentally acquired Maternal Engraftment) |
| **Leaky/atypical SCID** (low T cells) | Criteria 1 & 2 & 4  **OR** Criteria 1 & 3 & 4 | Two or more of:   - Low T-cell number for age (0.05-1.0 × 10^9^/L)[**#**](https://www.jacionline.org/article/S0091-6749(22)01479-8/fulltext) - Oligoclonal T cells****** - Abnormal TRECs OR <20% of CD4+ T cells are naive | At least a likely pathogenic variant | Reduced proliferation**††** | Does not have:   - Other SCID subtype - CID with known genotype - Thymic disorder - Other disorder with low T-cell numbers[**‡‡**](https://www.sciencedirect.com/science/article/pii/S0091674922014798#tbl1fnddaggerddagger) |
| **Omenn syndrome** | All 4 Criteria | >80% of CD4+ T cells have CD45RO+ memory phenotype | At least a likely pathogenic variant | Generalized rash  **AND** Absence of TME | Two or more of:   - Eosinophilia (>0.8 × 10^9^/L = >800μL) - Elevated IgE (1 reported upper limit of normal for children younger than 1 year is 34 IU/mL) - Abnormal TRECs - Lymphadenopathy - Hepatomegaly and/or splenomegaly - Oligoclonal T cells |

***** T-cell subset determination (with naive/memory phenotyping) should be repeated at least once, with the second test used as the criterion value. In patients with an identified pathogenic variant, the interval between tests must be at least 1 wk; however, in patients without an identified pathogenic gene variant, the T-cell number must remain <0.05 x 10^9^/L for at least 8 wk to qualify as typical SCID due to the potential for spontaneous improvement, with a shorter interval only if urgent hematopoietic cell transplant is required before 8 wk.

[**†**](https://www.jacionline.org/article/S0091-6749(22)01479-8/fulltext) At least a likely pathogenic variant identified in a gene whose product is known to be essential for T-cell development (examples in Table III, original publication^19^). Note that the pathogenic/likely pathogenic classification must be reached using the SCID VCEP specifications without criterion 2 to avoid circularity.

[**‡**](https://www.jacionline.org/article/S0091-6749(22)01479-8/fulltext) Alternate explanations for low T-cell counts include those listed in Criterion 4 of leaky/atypical SCID.

**§** Number of TRECs below the normal cutoff or cycle threshold value above the normal cutoff defined as consistent with SCID by performing laboratory.

[||](https://www.sciencedirect.com/science/article/pii/S0091674922014798#tbl1fnparpar)Naive T cells should be measured via CD3/CD4/CD45RA, or with additional naive markers.

[**¶**](https://www.jacionline.org/article/S0091-6749(22)01479-8/fulltext) Best performed by DNA analysis, such as with short tandem repeats, from whole blood or CD3-separated cells, with any level of detection considered positive. Documented TME classifies patients as typical SCID; TME testing is strongly recommended for patients considered to possibly have leaky/atypical SCID.

**#** Low T-cell numbers for age defined as <0.6 x 10^9^/L (any age), <0.8 x 10^9^/L if aged 2-4 y, or <1.0 x 10^9^/L if younger than 2 y.

****** Oligoclonal T cells as defined by laboratory performing testing, eg, <5 peaks in >4 T-cell receptor (TCR) Vbeta families on spectratyping, evidence of expansion of ≥2 TCR

Vbeta families to >2 X the upper limit of normal for those families, or low Shannon [H] entropy index on high-throughput sequencing of TCR Vbeta variable regions.

**††** Reduced proliferation is defined as a proliferative response to PHA, anti-CD3, or anti-CD3/CD28 <50% lower limit of reference range for laboratory.

[**‡‡**](https://www.sciencedirect.com/science/article/pii/S0091674922014798#tbl1fnddaggerddagger) Examples in Table II, original publication^19^.

Supplemental Table S2. Specifications for determining the strength of PS2/PM6

| **Phenotypic consistency** | **Points per proband** | |
| --- | --- | --- |
|  | ***de novo* with confirmed parental relationships** | ***de novo* with unconfirmed parental relationships** |
| Phenotype highly specific for gene | 2 | 1 |
| Phenotype consistent with gene but not highly specific | 1 | 0.5 |
| Phenotype consistent with gene but not highly specific and highly heterogeneity ^a^ | 0.5 | 0.25 |
| Phenotype not consistent with gene | 0 | 0 |
| **Strength of PS2 or PM6** | **The point total for all probands** | |
| Supporting | 0.5 | |
| Moderate | 1 | |
| Strong | 2 | |
| Very Strong | 4 | |

^a^ Maximum allowable value of 1 may contribute to the overall score

Supplemental Table S3. Specifications for determining the strength of PM3

| **Classification/Zygosity**  **of other variant ^a^** | **Points per proband** | |
| --- | --- | --- |
|  | **Confirmed in *trans*** | **Phase unknown** |
| Pathogenic or Likely Pathogenic variant | 1.0 | 0.5 (Pathogenic)  0.25 (Likely Pathogenic) |
| Homozygous occurrence  *(max point 1.0)* | 0.5 | N/A |
| Uncertain significance variant  *(max point 0.5)* | 0.25 | 0.0 |
| **Strength of PM3** | **The point total for all probands** | |
| Supporting | 0.5 | |
| Moderate | 1 | |
| Strong | 2 | |
| Very Strong | 4 | |

^a^All variants should be sufficiently rare (meet PM2 specification)

Supplemental Table S4. Pilot phase results

| **Variant Information** | **ClinVar ID** | **ClinVar classification** | **SCID VCEP classification** | **Codes applied by SCID VCEP** |
| --- | --- | --- | --- | --- |
| NM_000022.2(ADA):c.22G>A  (p.Asp8Asn) | 1973 | Benign | Benign | BA1 and BS2 |
| NM_000022.4(ADA):c.532del (p.Val177_Val178insTer) | 505549 | Pathogenic/  Likely pathogenic | Pathogenic | PVS1, PP4, PM2_Supporting, and PM3_Supporting |
| NM_000022.4(ADA):c.454C>A  (p.Leu152Met) | 1979 | Conflicting (Pathogenic,  Likely pathogenic, and VUS) | VUS | PP1_Supporting and PP4_Moderate |
| NM_000022.4(ADA):c.445C>T  (p.Arg149Trp) | 68264 | VUS | Likely pathogenic | PM2_Supporting, PP4_Moderate, and PS3_Moderate |
| NM_000022.4(ADA):c.446G>A  (p.Arg149Gln) | 1963 | VUS | VUS | BS3_Supporting and PM5_Supporting |
| NM_000022.4(ADA):c.703C>T  (p.Arg235Trp) | 468281 | Pathogenic/  Likely pathogenic | Likely pathogenic | PM2_Supporting, PP4, PM3_Strong, and PM5_Supporting |
| NM_000022.4(ADA):c.201C>G  (p.Tyr67Ter) | 1075328 | Pathogenic | Likely pathogenic | PVS1 and PM2_Supporting |
| NM_000022.4(ADA):c.632G>A  (p.Arg211His) | 1957 | Pathogenic/  Likely pathogenic | Pathogenic | PP4_Moderate, PM3_Strong, PP1_Strong, PM2_Supporting, and PS3_Moderate |
| NM_000022.4(ADA):c.396dup  (p.Val133fs) | 550821 | Pathogenic/  Likely pathogenic | Pathogenic | PVS1, PM2_Supporting, and PP4 |
| NM_000022.4(ADA):c.591T>A  (p.His197Gln) | 338509 | Conflicting  (Likely benign  and VUS) | Likely benign | BS1 |
| NM_000022.4(ADA):c.363-2A>G | 1473380 | Likely pathogenic | Likely pathogenic | PM2_Supporting, PVS1_Strong, and PP4 |
| NM_000022.4(ADA):c.219-2A>G | 1969 | Likely pathogenic | Likely pathogenic | PVS1_Strong, PM2_Supporting, and PM3_Supporting |
| NM_000022.4(ADA):c.402C>T  (p.Gly134=) | 536186 | Benign/  Likely benign | Likely benign | BS1, BS2_Supporting, and BP7 |
| NM_000022.4(ADA):c.192G>A  (p.Lys64=) | 719476 | Benign/  Likely benign | Likely benign | BS1 and BP7 |
| NM_001033855.3(DCLRE1C):c.1669dup  (p.Thr557fs) | 254216 | Pathogenic | Likely pathogenic | PVS1_Moderate, PM2_Supporting, PP1_Supporting, and PP4_Moderate |
| NM_001033855.3(DCLRE1C):c.406G>A (p.Asp136Asn) | 986350 | Conflicting  (Likely pathogenic  and VUS) | Likely pathogenic | PP4, PM3_Strong, PS3_Moderate, and PM2_Supporting |
| NM_001033855.3(DCLRE1C):c.959C>G (p.Ser320Cys) | 137073 | Benign | Benign | BA1 and BS2_Supporting |
| NM_001033855.3(DCLRE1C):c.512C>G (p.Pro171Arg) | 35999 | Benign | Benign | BA1 and BS2_Supporting |
| NM_001033855.3(DCLRE1C):c.346T>C (p.Cys116Arg) | 1713265 | Pathogenic | Likely pathogenic | PM2_Supporting, PP4_Strong, and PM3 |
| NM_001033855.3(DCLRE1C):c.47T>C (p.Ile16Thr) | 1069380 | Pathogenic | Likely pathogenic | PM3_Strong, PM2_Supporting, and PP4 |
| NM_001033855.3(DCLRE1C):c.1791C>T (p.Cys597=) | 771619 | Benign | Likely benign | BS1 and BP7 |
| NM_001033855.3(DCLRE1C):c.457G>A (p.Gly153Arg) | 35998 | Conflicting  (Benign, Likely benign, and VUS) | Benign | BA1 and BS2_Supporting |
| NM_001033855.3(DCLRE1C):c.194C>T (p.Thr65Ile) | 254217 | Pathogenic | Pathogenic | PM2_Supporting, PP4_Moderate, PP1_Strong, and PM3_Strong |
| NM_001033855.3(DCLRE1C):c.169G>T (p.Val57Phe) | 299322 | Conflicting  (Likely benign  and VUS) | Likely benign | BS1 |
| NM_001033855.3(DCLRE1C):c.973-1801T>A | 1679474 | Likely benign | VUS | BP7 |
| NM_001033855.3(DCLRE1C):c.247-1G>C | 1696158 | Likely pathogenic | VUS | PVS1_Moderate and PM2_Supporting |
| NM_002185.5(IL7R):c.265C>T  (p.Gln89Ter) | 804345 | Pathogenic/  Likely pathogenic | Likely pathogenic | PVS1 and PM2_Supporting |
| NM_002185.5(IL7R):c.1241C>T (p.Thr414Met) | 134536 | Benign | Benign | BA1 and BS2_Supporting |
| NM_002185.5(IL7R):c.83-2A>T | 353259 | Conflicting  (Pathogenic  and VUS) | Pathogenic | PVS1, PM2_Supporting, and PM3_Supporting |
| NM_002185.5(IL7R):c.353G>A  (p.Cys118Tyr) | 36392 | Pathogenic/  Likely pathogenic | Pathogenic | PM2_Supporting, PM3_VeryStrong, PP1, and PP4_Moderate |
| NM_002185.5(IL7R):c.361dup  (p.Ile121fs) | 224841 | Pathogenic/  Likely pathogenic | Pathogenic | PVS1, PM2_Supporting, and PP4. |
| NM_002185.5(IL7R):c.662G>T  (p.Ser221Ile) | 134528 | Pathogenic/  Likely pathogenic | Pathogenic | PM2_Supporting, PM3_VeryStrong, and PP4. |
| NM_002185.5(IL7R):c.704C>G  (p.Ser235Ter) | 578174 | Pathogenic | Likely pathogenic | PVS1 and PM2_Supporting |
| NM_002185.5(IL7R):c.132C>T  (p.Ser44=) | 377976 | Benign/  Likely benign | Likely benign | BP7 and BS1 |
| NM_002185.5(IL7R):c.1231A>G (p.Thr411Ala) | 134534 | Benign/  Likely benign | Likely benign | BS1 and BS2_Supporting |
| NM_002185.5(IL7R):c.778G>A  (p.Ala260Thr) | 418258 | Conflicting  (Likely benign  and VUS) | Likely benign | BS1 |
| NM_002185.5(IL7R):c.495C>T  (p.His165=) | 137585 | Benign | Benign | BA1, BS2_Supporting, and BP7 |
| NM_002185.5(IL7R):c.1020T>G  (p.Leu340=) | 709483 | Benign/  Likely benign | Likely benign | BS1, BS2_supporting, and BP7 |
| NM_002185.5(IL7R):c.82+14A>T | 2187538 | Likely benign | VUS | PM2_supporting, and BP7 |
| NM_002185.5(IL7R):c.538-1G>A | 14841 | Pathogenic | Pathogenic | PM2_Supporting, PVS1, PP4, and PM3 |
| NM_000448.3(RAG1):c.2487_2488delinsTT (p.Arg829_Lys830delinsSerTer) | 1034220 | Conflicting  (Pathogenic  and VUS) | Pathogenic | PVS1, PM2_Supporting, PM3, and PP4 |
| NM_000448.3(RAG1):c.1A>G  (p.Met1Val) | 304491 | Conflicting  (Likely pathogenic  and VUS) | VUS | PVS1_Moderate and PM2_Supporting |
| NM_000448.3(RAG1):c.256_257del (p.Lys86fs) | 285045 | Pathogenic | Pathogenic | PVS1, PM2_Supporting, PM3_Moderate, PS3_Moderate, and PP4_Supporting |
| NM_000448.3(RAG1):c.527G>T (p.Cys176Phe) | 372487 | Likely pathogenic | VUS | PM2_Supporting, PM3, and PP4 |
| NM_000448.3(RAG1):c.577G>A (p.Glu193Lys) | 304495 | Benign | Benign | BA1 and BS2_Supporting |
| NM_000448.3(RAG1):c.725A>G (p.Gln242Arg) | 536967 | Conflicting  (Benign  and VUS) | Likely benign | BS1 |
| NM_000448.3(RAG1):c.906C>A (p.Asp302Glu) | 36715 | Benign/  Likely benign | Benign | BA1 |
| NM_000448.3(RAG1):c.424C>T  (p.Arg142Ter) | 626157 | Pathogenic/  Likely pathogenic | Pathogenic | PVS1, PM3, and PM2_Supporting |
| NM_000448.3(RAG1):c.775del  (p.Ser259fs) | 235411 | Pathogenic | Pathogenic | PVS1, PM3_Supporting, and PM2_Supporting |
| NM_000448.3(RAG1):c.1331C>T (p.Ala444Val) | 68681 | Pathogenic | Pathogenic | PM1, PM2_Supporting, PP4, PM3_Strong, and PS3_Moderate |
| NM_000448.3(RAG1):c.2751G>A  (p.Gln917=) | 304506 | Benign/  Likely benign | Likely benign | BS1 and BP7 |
| NM_000448.3(RAG1):c.303G>A  (p.Ala101=) | 138882 | Benign | Benign | BA1, BS2_supporting, and BP7 |
| NM_000536.4(RAG2):c.1357T>A (p.Trp453Arg) | 496630 | Pathogenic/  Likely pathogenic | Likely pathogenic | PM1, PS3_Moderate, PP4, PM2_Supporting, and PM3_Supporting |
| NM_000536.4(RAG2):c.283G>A  (p.Gly95Arg) | 13133 | Pathogenic/  Likely pathogenic | Likely pathogenic | PM1_Supporting, PM2_Supporting, PS3_Moderate, PP4, and PM3 |
| NM_000536.4(RAG2):c.829dup  (p.Tyr277fs) | 1075544 | Pathogenic | Pathogenic | PVS1, PM2_Supporting, PS3_Moderate, and PP4 |
| NM_000536.4(RAG2):c.1095T>C  (p.Ser365=) | 304552 | Benign/  Likely benign | Likely benign | BS1 and BP7 |
| NM_000536.4(RAG2):c.686G>A (p.Arg229Gln) | 13130 | Conflicting  (Pathogenic,  Likely pathogenic  and VUS) | Pathogenic | PM2_Supporting, PP4, PM3_Moderate, PS3, PP1, and PM1_Supporting |
| NM_000536.4(RAG2):c.955G>T  (p.Gly319Ter) | 500475 | Pathogenic | Likely pathogenic | PVS1 and PM2_Supporting |
| NM_000536.4(RAG2):c.217C>T  (p.Arg73Cys) | 36719 | Pathogenic/  Likely pathogenic | Likely pathogenic | PP4, PM1_Supporting, PM3, PM5_Supporting, and PM2_Supporting |
| NM_000536.4(RAG2):c.1158C>A (p.Phe386Leu) | 138887 | Benign/  Likely benign | Benign | BA1 and BS2_Supporting |
| NM_000536.4(RAG2):c.22G>A  (p.Val8Ile) | 138885 | Benign/  Likely benign | Likely benign | BS1 and BS2_Supporting |
| NM_000536.4(RAG2):c.130G>T  (p.Gly44Ter) | 1412375 | Pathogenic | Pathogenic | PVS1, PM2_Supporting, PP4, and PM3_Supporting |
| NM_000536.4(RAG2):c.1352G>C (p.Gly451Ala) | 13138 | Conflicting (Pathogenic,  Likely pathogenic  and VUS) | Likely pathogenic | PM2_Supporting, PM1, PS3_Supporting, and PM3 |
| NM_000536.4(RAG2):c.193G>T  (p.Asp65Tyr) | 427020 | Conflicting (Pathogenic,  Likely pathogenic  and VUS) | Likely pathogenic | PM2_Supporting, PS3_Moderate, PM1_Supporting, PM3_Supporting, and PP4 |
| NM_000215.4(JAK3):c.1796T>G (p.Val599Gly) | 624608 | NA | VUS | PM2_Supporting and PP4_Moderate |
| NM_000215.4(JAK3):c.1744C>T (p.Arg582Trp) | 36415 | Pathogenic/  Likely pathogenic | Likely pathogenic | PM2_Supporting, PM3, PM4_Supporting, and PP4_Moderate |
| NM_000215.4(JAK3):c.2625C>T  (p.Leu875=) | 36419 | Benign | Benign | BA1, BS2_Supporting, and BP7 |
| NM_000215.4(JAK3):c.2773C>A (p.Arg925Ser) | 418267 | Conflicting  (Likely pathogenic, VUS and  Likely benign) | Likely benign | BS1 |
| NM_000215.4(JAK3):c.896T>C  (p.Val299Ala) | 891294 | Conflicting  (Likely benign  and VUS) | VUS | PM2_Supporting |
| NM_000215.4(JAK3):c.1351C>T (p.Arg451Ter) | 81020 | Pathogenic | Pathogenic | PVS1, PM3, PP4, and PM2_Supporting |
| NM_000215.4(JAK3):c.678_679del (p.Cys227fs) | 36423 | Pathogenic/  Likely pathogenic | Pathogenic | PVS1, PP4, PM2_Supporting, and PM3_Supporting |
| NM_000215.4(JAK3):c.1837C>T (p.Arg613Ter) | 644288 | Pathogenic | Pathogenic | PVS1, PP4_Moderate, PM2_Supporting, and PM3_Supporting |
| NM_000215.4(JAK3):c.2636A>G (p.His879Arg) | 372390 | Conflicting  (Likely benign  and VUS) | Likely benign | BS1 |
| NM_000215.4(JAK3):c.175A>T  (p.Lys59Ter) | 1999662 | Pathogenic | Likely pathogenic | PVS1 and PM2_Supporting |
| NM_000215.4(JAK3):c.1915-1G>A | 2048620 | Likely pathogenic | VUS | PM2_Supporting |
| NM_000215.4(JAK3):c.2680+89G>A | 2054022 | Pathogenic | Likely pathogenic | PM2_supporting, PP1_Strong, PM3_Supporting, PP4_Moderate, and PP3. |
| NM_000206.3(IL2RG):c.924G>A  (p.Ser308=) | 962267 | Conflicting  (Pathogenic  and VUS) | Likely pathogenic | PM2_Supporting, PP3, PP4_Moderate, PP1, and PM4 |
| NM_000206.3(IL2RG):c.43C>T  (p.Gln15Ter) | 372386 | Pathogenic/  Likely pathogenic | Pathogenic | PVS1, PM2_Supporting, and PP4 |
| NM_000206.3(IL2RG):c.332T>C  (p.Ile111Thr) | 795305 | Benign | Likely benign | BS2 |
| NM_000206.3(IL2RG):c.924+1G>A | 280035 | Pathogenic | Pathogenic | PVS1, PM2_Supporting, and PP4_Moderate |
| NM_000206.3(IL2RG):c.977G>A (p.Ser326Asn) | 1166432 | Benign | Likely benign | BS2 |
| NM_000206.3(IL2RG):c.325G>A (p.Glu109Lys) | 368619 | Benign/  Likely benign | Benign | BS1 and BS2 |
| NM_000206.3(IL2RG):c.676C>T (p.Arg226Cys) | 225195 | Pathogenic | Pathogenic | PM2_Supporting, PM1_Strong, PM6_Supporting, PS4, PP4, PS3_Supporting |
| NM_000206.3(IL2RG):c.664C>T (p.Arg222Cys) | 10027 | Pathogenic | Pathogenic | PM2_Supporting, PP1_Strong, PS4, and PP4_Moderate |
| NM_000206.3(IL2RG):c.1105A>G (p.Thr369Ala) | 837417 | Likely benign | VUS | BS2_Supporting |
| NM_000206.3(IL2RG):c.821T>C  (p.Ile274Thr) | 532191 | VUS | VUS | PM2_Supporting and PM1_Strong |
| NM_000206.3(IL2RG):c.963G>A  (p.Leu321=) | 763173 | Benign | Likely benign | BS2 and BP7 |
| NM_000206.3(IL2RG):c.924+9G>T | 1559662 | Likely benign | VUS | PM2_Supporting and BP7 |
| NM_000206.3(IL2RG):c.115+2T>C | 1368945 | Pathogenic | Pathogenic | PVS1, PM2_Supporting, and PP4 |
| NM_000206.3(IL2RG):c.694G>A (p.Gly232Arg) | 633274 | NA | VUS | PM2_Supporting and PP4 |
